# Increased Insular Functional Connectivity During Repetitive Negative Thinking in Major Depression and Healthy Volunteers

**DOI:** 10.1101/2024.10.15.24315550

**Authors:** Landon S Edwards, Saampras Ganesan, Jolene Tay, Eli S Elliott, Masaya Misaki, Evan J White, Martin P Paulus, Salvador M Guinjoan, Aki Tsuchiyagaito

**Affiliations:** Laureate Institute for Brain Research, Tulsa, OK, USA; Department of Biomedical Engineering, The University of Melbourne, Carlton, Victoria 3053, Australia; Contemplative Studies Centre, Melbourne School of Psychological Sciences, The University of Melbourne, Melbourne, Victoria 3010, Australia; Oxley College of Health and Natural Sciences, The University of Tulsa, Tulsa, OK, USA; Department of Psychiatry, Oklahoma University Health Sciences Center at Tulsa, Tulsa, OK, USA; Laureate Psychiatric Hospital and Clinic, Tulsa, OK, USA; Research Center for Child Mental Development, Chiba University, Chiba, Japan

**Keywords:** insula, repetitive negative thinking, rumination, depression, functional connectivity

## Abstract

**Background:** Repetitive negative thinking (RNT) in major depressive disorder (MDD) involves persistent focus on negative self-related experiences. Resting-state fMRI shows that the functional connectivity (FC) between the insula and the superior temporal sulcus is critical to RNT intensity. This study examines how insular FC patterns differ between resting-state and RNT-induction in MDD and healthy participants (HC).

**Methods:** Forty-one individuals with MDD and twenty-eight HCs (total n=69) underwent resting-state and RNT-induction fMRI scans. Seed-to-whole brain analysis using insular subregions as seeds was performed.

**Results:** No diagnosis-by-run interaction effects were observed across insular subregions. MDD participants showed greater FC between bilateral anterior, middle, and posterior insular regions and the cerebellum (z = 4.31 to 6.15). During RNT-induction, both MDD and HC participants demonstrated increased FC between bilateral anterior and middle insula and key brain regions, including prefrontal cortices, parietal lobes, posterior cingulate cortex, and medial temporal gyrus, encompassing the STS (z = 4.47 to 8.31). Higher trait-RNT was associated with increased FC between the right dorsal anterior and middle insula and regions in the DMN and salience network in MDD participants (z = 4.31 to 6.15). Greater state-RNT scores were linked to increased FC in similar insular regions, the bilateral angular gyrus and right middle temporal gyrus (z = 4.47 to 8.31).

**Conclusions:** Hyperconnectivity in insula subregions during active rumination, especially involving the DMN and salience network, supports theories of heightened self-focused and negative emotional processing in depression. These findings emphasize the neural basis of RNT when actively elicited in MDD.

## 1. Introduction

Repetitive Negative Thinking (RNT), such as rumination in the context of depression, is a cognitive process characterized by a persistent focus on negative experiences related to the self (Nolen-Hoeksema *et al*., 2008). RNT is a symptom dimension with significant implications for the course and prognosis of depression, making this disorder refractory to treatment, chronic, and complicated with suicide (Krajniak *et al*., 2013, Surrence *et al*., 2009, Watkins and Roberts, 2020). Previous research has examined the triggers, intensity, and duration of RNT. Characterizing the neurobiological mechanisms of RNT is important not only for understanding its formation, but also to discern targets for neuromodulation addressed at alleviating this symptom.

Prior functional connectivity-based (FC) studies have identified many regions of interest (ROIs) as they relate to heightened RNT and brooding symptoms in individuals, including the left dorsolateral prefrontal cortex, precuneus, and other components of the default mode network (DMN) (Jacob *et al*., 2020, Taylor *et al*., 2022). However, our previous resting-state fMRI study revealed that RNT intensity correlates with increased FC between the bilateral anterior insular cortices and the right superior temporal sulcus (STS) (Tsuchiyagaito *et al*., 2022). This result highlighted the neural mechanisms underlying RNT as difficulties in disengaging attention from negative emotional responses (Craig, 2009), and having interrelation with inner-speech processing (Deen *et al*., 2015). This is compatible with the view that the DMN serves resting self-dialogue, but not necessarily depressive rumination (Goldstein-Piekarski *et al*., 2022). Thus, prior evidence deemphasizes the role of DMN dysfunction in RNT (Goldstein-Piekarski *et al*., 2022, Makovac *et al*., 2020, Tozzi *et al*., 2021), while recent work by our group (Tsuchiyagaito *et al*., 2022) demonstrates that the functional connection between the insula (Craig, 2009) and the STS (Deen *et al*., 2015) is related to the intensity of RNT (Tsuchiyagaito *et al*., 2022). Nevertheless, our understanding is limited to the resting-state data, which lacks clarity on the RNT circuit when individuals are actively engaging with RNT.

RNT has been established as a trait-like cognitive process which involves recurrent and continuous focus on self-relevant negative thoughts that is persistent over time and across situations. However, RNT intensity can also fluctuate, such that there is a state component to it; it can be influenced by overall depression symptom severity, instant mood state, and adverse environmental stimuli - including relevant interpersonal interactions (Chang *et al*., 2023, Philippi *et al*., 2022). This differentiation aligns with recent studies utilizing the experimental induction of RNT, which demonstrates the potential independence and distinct characteristics of both trait- and state-RNT (Grant *et al*., 2021, LeMoult *et al*., 2013, Robinson and Alloy, 2003, Wang *et al*., 2022). For example, Misaki *et al*. (2023) highlighted that while RSFC alterations distinguish between healthy and depressed individuals, trait-RNT in depressed individuals is more closely predicted by functional connectivity during an induced RNT scan rather than resting-state scan, suggesting that RNT involves an active mental process not fully represented in the resting-state. While trait-RNT measures an individual’s tendency to engage in RNT, induced RNT (capturing instant symptomatology) enables us to probe for specific triggers, response patterns, and the phenomenological characteristics of RNT that are not captured by trait-RNT alone. Thus, discerning the brain mechanisms that underlie both the trait and state aspects of RNT could have significant implications for clinical practice in terms of RNT remediation.

Given the results of our previous resting-state FC investigations and the prior literature, we aimed to further clarify the mechanistic basis of RNT by comparing insular FC during RNT-induction with resting-state FC in individuals with MDD. Specifically, we employed a seed-to-whole-brain analysis using six insula subregions as seeds. We hypothesized that individuals with MDD would exhibit a more substantial increase in insular FC during RNT-induction compared to resting-state, with these alterations being more pronounced in the MDD individuals than in HC. By investigating these neural dynamics, we seek to address the question: how do the functional connectivity patterns of the insula differ between resting-state and RNT-induction in MDD, and what implications do these differences have for the development of targeted neuromodulatory interventions?

## 2. Methods

### 2.1 Study Design

The study protocol was reviewed and approved by the WCG IRB (https://www.wcgirb.com) (IRB Tracking Number 20210286), and registered on ClinicalTrials.gov (NCT04941066) as a part of a real-time fMRI-neurofeedback (rtfMRI-nf) study (Tsuchiyagaito *et al*., 2023b, Tsuchiyagaito *et al*., 2021).

### 2.2 Participants

Forty-one MDD and twenty-eight healthy control (HC) volunteers were recruited for rtfMRI-nf studies, making up a total of sixty-nine participants (Tsuchiyagaito *et al*., 2023b, Tsuchiyagaito *et al*., 2021). Participants were of both sexes, between the ages of 18 and 65 years old, and fluent in English. Exclusion criteria were pregnancy, an abnormal neuromorphological brain profile as assessed by a radiology specialist physician, and other general contraindications for MRI safety. HC participants were defined based on the Mini-International Neuropsychiatric Interview 7.0.2 (MINI) (Sheehan *et al*., 1998), and confirmed in a clinical conference with a board-certified psychiatrist. MDD-specific inclusion criteria included: meeting the criteria of the 5th edition of the *Diagnostic and Statistical Manual of Mental Disorders* (DSM-5) for unipolar MDD based on the MINI (Sheehan *et al*., 1998) and current depressive symptoms with a Montgomery-Åsberg Depression Rating Scale (MADRS) score of > 6 (Montgomery and Asberg, 1979). MDD-specific exclusion criteria were as follows: a lifetime history of bipolar disorder, schizophrenia, or any psychotic disorders; DSM-5 criteria for substance abuse or dependence within six months prior to study entry; active suicidal ideation as indicated by the Columbia-Suicide Severity Rating Scale (C-SSRS) (Posner *et al*., 2011) or an attempt within 12 months prior to study entry; commencement of psychotropic medication for depression and/or anxiety less than one month before the study enrollment; commencement of psychological therapy less than one month before the study enrollment. All participants completed a written, informed consent process before participating in the study.

### 2.3 Neuroimaging data acquisition

Neuroimaging was conducted on a 3 Tesla MR750 Discovery scanner (GE Healthcare, Milwaukee, WI) with an 8-channel, receive-only head array coil. Blood-oxygen-level-dependent fMRI data were acquired using a T2*-weighted gradient echo-planar sequence with sensitivity encoding (GE-EPI SENSE) with the following parameters: TR/TE = 2000/25 ms, acquisition matrix = 96 × 96, FOV/slice = 240/2.9 mm, flip angle = 90°, voxel size = 2.5×2.5×2.9 mm; 40 axial slices, SENSE acceleration R = 2. To provide anatomical reference for fMRI data, T1-weighted (T1w) MRI images were acquired with a magnetization-prepared rapid gradient-echo (MPRAGE) sequence with the parameters of FOV = 240×192 mm, matrix = 256×256, 124 axial slices, slice thickness = 1.2 mm, 0.94×0.94×1.2 mm^3^ voxel volume, TR/TE = 5/2 ms, SENSE acceleration R = 2, flip angle = 8°, delay/inversion time TD/TI = 1400/725 ms, sampling bandwidth = 31.2 kHz, scan time = 4 min 59 s.

### 2.4 Experimentally induced RNT and resting-state scanning

The MRI session started with a 5 min T1w MRI anatomical scan, 6 min 50 s resting-state fMRI scan, and a 6 min 50 s experimentally induced RNT fMRI scan. Prior to the MRI session, participants identified a recent personal event that significantly triggered RNT, such as experiencing rejection by someone important to them. Participants provided a brief title for this event, which was used by research staff to prompt the participant’s recall immediately before the RNT-inducing fMRI scan. Participants were then instructed about the neurofeedback task as described in detail in Tsuchiyagaito *et al*. (2023b), Tsuchiyagaito *et al*. (2021), and then had a rest period before the MRI session. In the scanner, the session began with a resting-state scan, where participants were instructed to clear their mind and not think of anything while viewing a fixation cross. This was followed by the RNT-inducing fMRI scan, during which participants were reminded of their chosen event and instructed to introspectively ruminate and ponder on it. While keeping their gaze on the fixation cross, they were asked to focus on their emotional reactions to their chosen event and why they responded the way they did. This procedure aimed to engage the participants in a state of rumination and brooding, characteristic of RNT, while inside the scanner. The MRI session ended with neurofeedback scans as described elsewhere (Tsuchiyagaito *et al*., 2023b, Tsuchiyagaito *et al*., 2021).

### 2.5 Symptom measures Trait-RNT

The 22-item Ruminative Response Scale (RRS) (Nolen-Hoeksema and Morrow, 1991) was used to measure trait-RNT. The RRS is composed of three subscales; the 5-item ’brooding’ subscale (e.g., RRS-B item: *think* ‘*why can’t I handle things better’*), the 12-item ‘depressive rumination’ subscale (e.g., RRS-D item: *think about all of your shortcomings, failings, faults, and mistakes*), and the 5-item ‘reflection’ subscale (e.g., RRS-R item: *write down what you are thinking and analyse it*). It assesses an individual’s tendency or trait to ruminate when they feel sad or are faced with depressive symptoms. Participants are asked to indicate what they “generally do when feeling down, sad, or depressed” using a 4-point Likert scale ranging from 1 (never) to 4 (always), representing the trait tendency. The items in the RRS-B measure how often people engage in RNT, the causes and consequences of RNT, or a passive comparison with unachieved goals – characteristics that are found to lead to worse prognoses of depression (Treynor *et al*., 2003). The items in the RRS-D subscale are similar to the RRS-B subscale; however, this subscale measures how often people engage in RNT, with a focus on the depressive symptoms and moods. We employed the RRS-B and RRS-D subscales for connectivity analyses related to trait-RNT. The RRS-R subscale was not included in our main analysis (results are shown in the Supplementary materials 2.1) as it does not include pathological elements of RNT and may even reflect protective factors against depression (Treynor *et al*., 2003).

#### State-RNT

The level of state-RNT that immediately followed the state-RNT fMRI scan was assessed with the visual analogue scale (VAS). Right after the resting-state and RNT-induction scans, participants used a button box to answer the question, *“To what extent did you dwell on negative aspects of yourself?”*. The answers consisted of ratings from 1 (not at all) to 10 (extremely), indicating the intensity of their state-RNT during the scan.

#### Severity of depression and anxiety

Individuals with MDD were assessed before the MRI session using the Montgomery-Åsberg Depression Rating Scale (MADRS) (Montgomery and Asberg, 1979) and the Hamilton Anxiety Scale (HAMA) (Maier *et al*., 1988).

### 2.6 Preprocessing

Preprocessing of functional images was performed with Analysis of Functional NeuroImages (AFNI) (http://afni.nimh.nih.gov/afni/). The initial three volumes were excluded from the analysis. The preprocessing included despiking, RETROICOR (Glover *et al*., 2000), respiratory volume per time (Birn *et al*., 2008) physiological noise corrections, slice-timing correction, motion corrections, nonlinear warping to the MNI template brain with resampling to 2 mm^3^ voxels using the Advanced Normalization Tools (Avants *et al*., 2008) (http://stnava.github.io/ANTs/), smoothing with 6mm-FWHM kernel, and scaling to percent change relative to the mean signal in each voxel. We used FastSurfer (https://www.sciencedirect.com/science/article/pii/S1053811920304985) to extract white matter and ventricle masks from the anatomical image of an individual subject and then warped them to the normalized fMRI image space. General linear model (GLM) analysis was performed with regressors of 12 motion parameters (three rotations, three shifts, and their temporal derivatives), three principal components of ventricle signals, local white matter average signals (ANATICOR (Jo *et al*., 2010)), 4th-order Legendre polynomials for high-pass filtering, and censoring TRs with large head motion (> 0.25 mm frame-wise displacement). Any data with more than 30% censored volumes was treated as a missing value for the group-level analysis (two datasets of HC during RNT-induction, and two datasets of MDD participants during RNT-induction and resting-state scans were treated as missing values). Voxel-wise residual signals of the GLM were used for the seed-to-whole brain analysis.

### 2.7 Seed-to-whole brain analysis Definition of insular subregions

In order to better delineate the specific function of the insula, the Brainnetome insula sub-regions parcellation atlas was used (Fan *et al*., 2016). This parcellation atlas defined fine-grained insular subregions using probabilistic connectivity patterns. The insula was segmented into six subregions in each hemisphere, including the hypergranular insula (G), ventral agranular insula (vla), dorsal agranular insula (dla) (Sliz and Hayley, 2012), ventral dysgranular and granular insula (vId/vIg), dorsal granular insula (dIg), and dorsal dysgranular insula (dId) (Supplementary materials, Figure S1).

#### FC processing

Twelve seed-to-whole brain FC maps were calculated based on predefined insular subregions. The average time-course was obtained from the seeds, and the FC maps were generated by calculating Pearson’s correlation coefficients between the time series within the seed and the time series from every other voxel across the whole brain. Correlation coefficients were converted to z-scores using Fisher’s r-to-z transformation.

#### Statistical analysis

AFNI’s 3dLMEr was performed on each seed to identify the connectivity patterns of the insular subregions with the interaction of diagnosis (MDD vs. HC) by run (RNT-induction vs. Rest), age, sex, motion, and medication status as fixed effects, and subjects as random intercepts. Results of the main interaction effect, main effect of diagnosis, and main effect of run were reported as a chi-square statistic, and post-hoc general linear t-style tests (GLT) were specified in case of the significant main effect, as per the output of AFNI’s 3dLMEr. The significant threshold was set as peak p < 0.001 and cluster-wise p < 0.05/12 (Bonferroni-corrected). AFNI’s 3dClustSim with 10,000 permutation tests were employed to define the cluster-size thresholds (k > 143 voxels). Furthermore, linear correlation analyses were performed to investigate the association between changes in FC values during RNT-induction scans compared to resting-state scans, and the trait- and Δstate-RNT (changes in RNT-induction relative to the baseline resting-state) in the MDD and HC groups, respectively. The uncorrected threshold p < 0.05 was considered significant for this exploratory correlation analysis.

## 3. Results

### 3.1 Demographic and clinical measures

Table 1 shows the demographic data and clinical characteristics of the MDD (n=41) and HC (n=28) participants (total n=69). The majority of these participants were Female and White, and over half of the MDD participants experienced anxiety disorder comorbidities (51.2%) and were treated with antidepressants (51.2%) (Table 1).

**Table 1.** Demographic data.

| Demographic data | MDD |  | HC |  |
| --- | --- | --- | --- | --- |
|  | Mean/Number | SD/% | Mean/Number | SD/% |
| Age | 36.54 | (12.27) | 23.04 | (3.92) |
| Female (%) | 31 | (75.61) | 21 | (75) |
| Handedness: Right (%) | 37 | (90.24) | 28 | (100) |
| Race/Ethnicity: Non-white (%) | 8 | (19.51) | 13 | (46.43) |
| Asian | 0 | (0) | 1 | (3.57) |
| Black/African American | 0 | (0) | 1 | (3.57) |
| Native Hawaiian/Pacific Islander | 0 | (0) | 1 | (3.57) |
| Hispanic | 1 | (2.44) | 7 | (25) |
| Indigenous | 7 | (17.07) | 1 | (3.57) |
| Multiracial | 0 | (0) | 2 | (7.14) |
| White | 33 | (80.49) | 15 | (53.57) |
| Diagnosis (%) |  |  |  |  |
| Major depressive disorder (MDD) without comorbidity | 20 | (48.78) |  |  |
| MDD and anxiety disorder | 21 | (51.22) |  |  |
| Generalized anxiety disorder | 13 | (31.71) |  |  |
| Social anxiety disorder | 7 | (17.07) |  |  |
| Panic disorder | 10 | (24.39) |  |  |
| Depressive episode (%) |  |  |  |  |
| Single episode | 14 | (34.15) |  |  |
| Recurrent | 27 | (65.85) |  |  |
| Medicated (%) | 21 | (51.22) |  |  |
| Antidepressants | 18 | (43.9) |  |  |
| Stimulants | 5 | (12.2) |  |  |
| Sedatives | 7 | (17.07) |  |  |
| Current psychotherapy (%) | 9 | (21.95) |  |  |
| Clinical measures |  |  |  |  |
| RRS Brooding | 12.61 | (3.13) | 6.46 | (1.53) |
| RRS Depressive rumination | 30.76 | (6.76) | 15.36 | (3.96) |
| RRS Reflection | 11.07 | (3.34) | 6.79 | (3.13) |
| RRS Total | 54.44 | (11.60) | 28.61 | (7.28) |
| State-RNT (VAS) - Rest | 2.59 | (2.23) | 2.03 | (1.28) |
| State-RNT (VAS) - Induction | 7.08 | (1.66) | 5.61 | (1.81) |
| MADRS | 19.9 | (5.86) |  |  |
| HAMA | 16.15 | (5.22) |  |  |

### 3.2 Insular-to-whole brain FC patters Interaction effect of diagnosis-by-run

We first examined the interaction effect of diagnosis (MDD vs HC) by run (resting-state vs RNT-induction). Contrary to our hypothesis, no significant FC alterations were observed for the diagnosis-by-run interaction across any of the insular subregions. Results with a threshold of p < 0.001, without cluster thresholding, are presented in the **Supplementary Materials, Figures S2 and S3.**

#### Main effect of diagnosis and run

Participants with MDD demonstrated greater FC between the bilateral anterior, middle, and posterior insular regions and the cerebellum (z = 4.31–6.15). These results suggest a unique pattern of insular-cerebellar connectivity in MDD (**Table 2**, and **Supplementary Figures S4 and S5**).

**Table 2.**
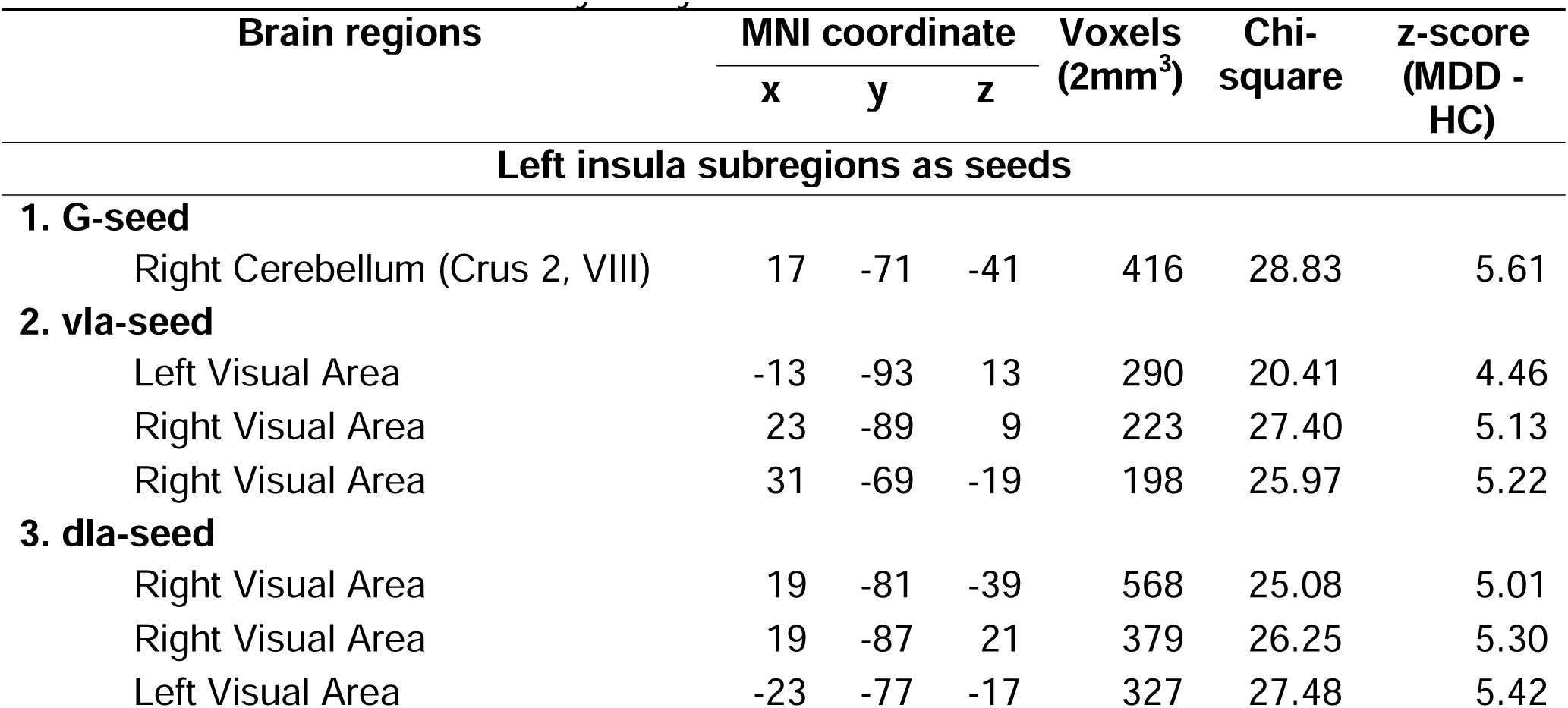
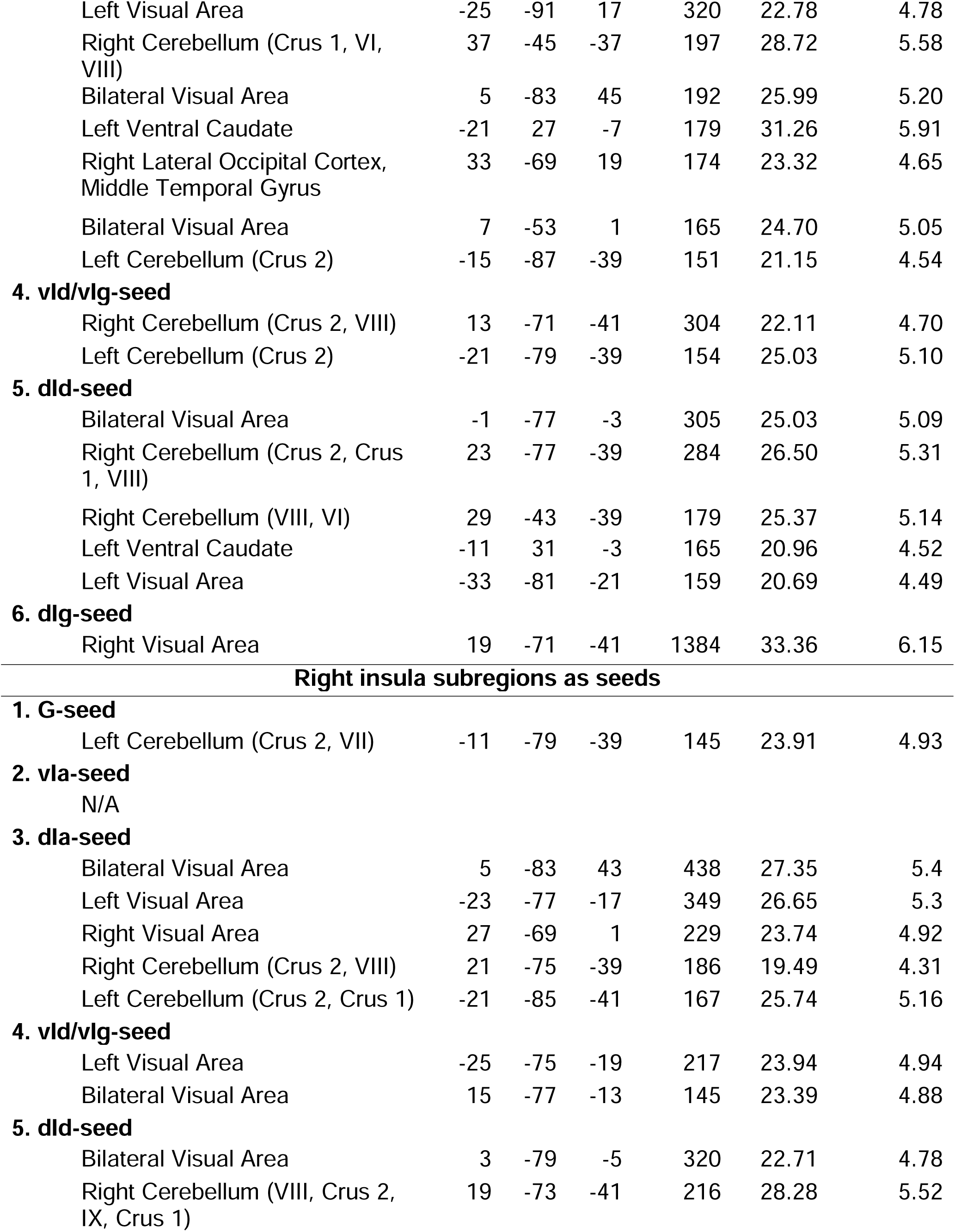
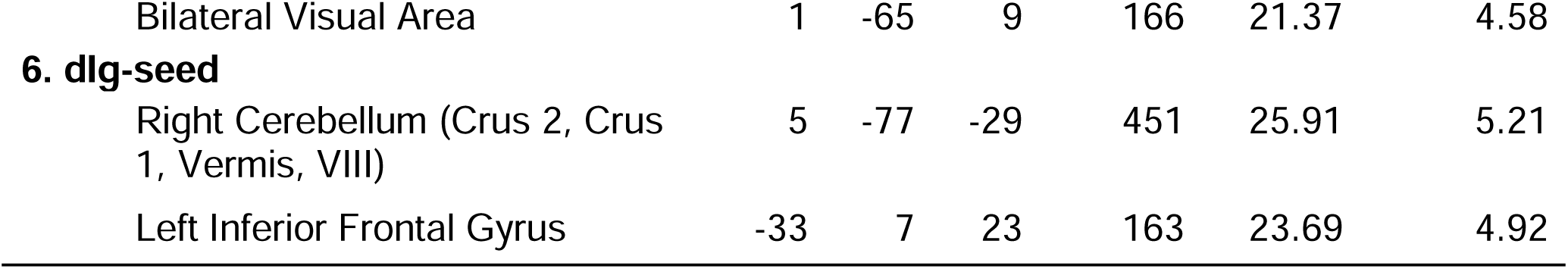
Significant regions showing main effect of diagnosis (MDD and HC) from seed-to-whole brain functional connectivity analysis.

Regarding the main effect of run (**Table 3**, and **Supplementary Figures S6 and S7**), enhanced FC was found between the bilateral anterior and middle insula and other key brain regions, including the bilateral prefrontal cortices, parietal lobes, posterior cingulate cortex, and medial temporal gyrus, encompassing the STS (z = 4.47–8.31). Figure 1 displays additional spider charts and bar plots to illustrate the post-hoc effects of these main findings.

**Table 3.**
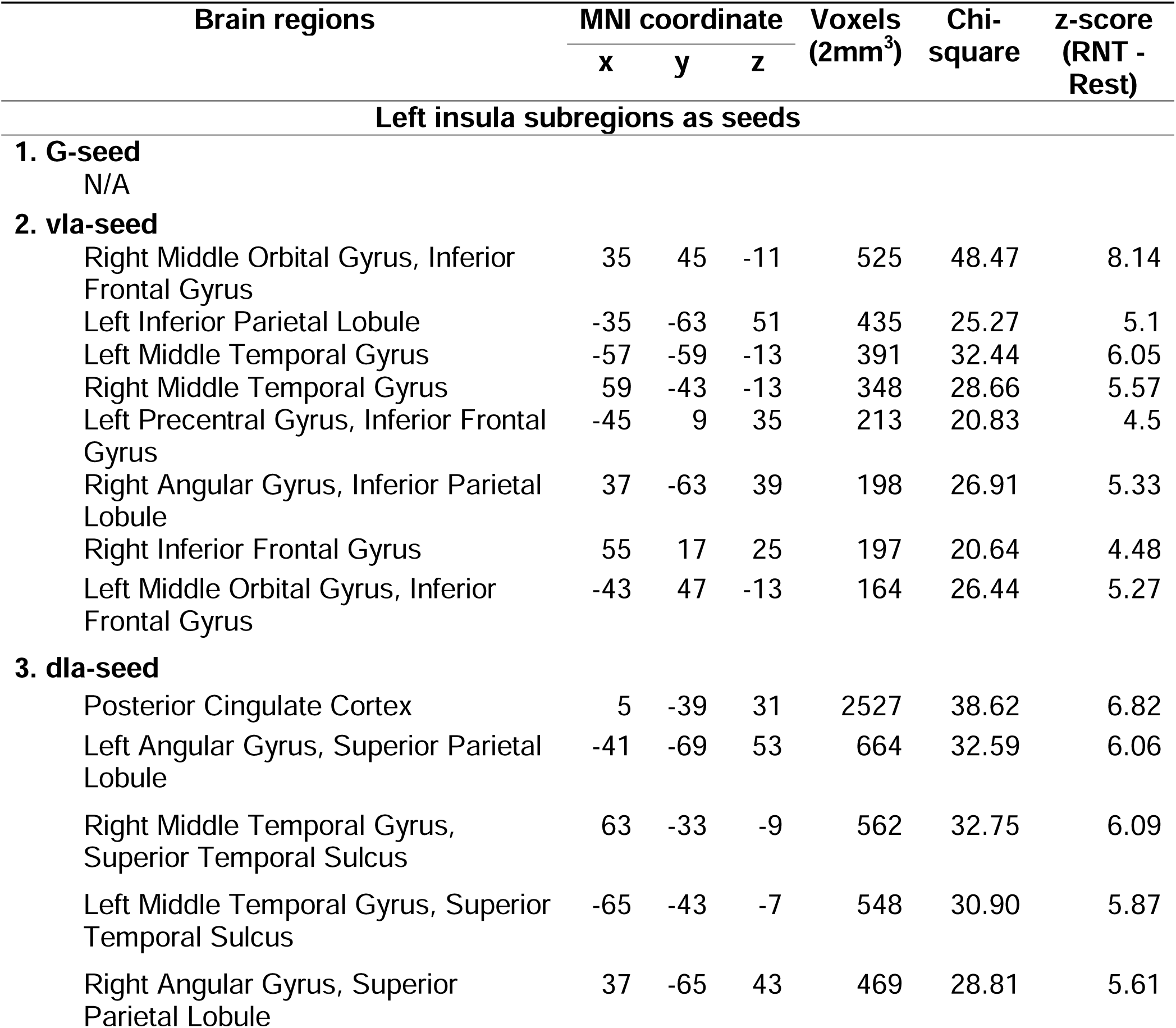
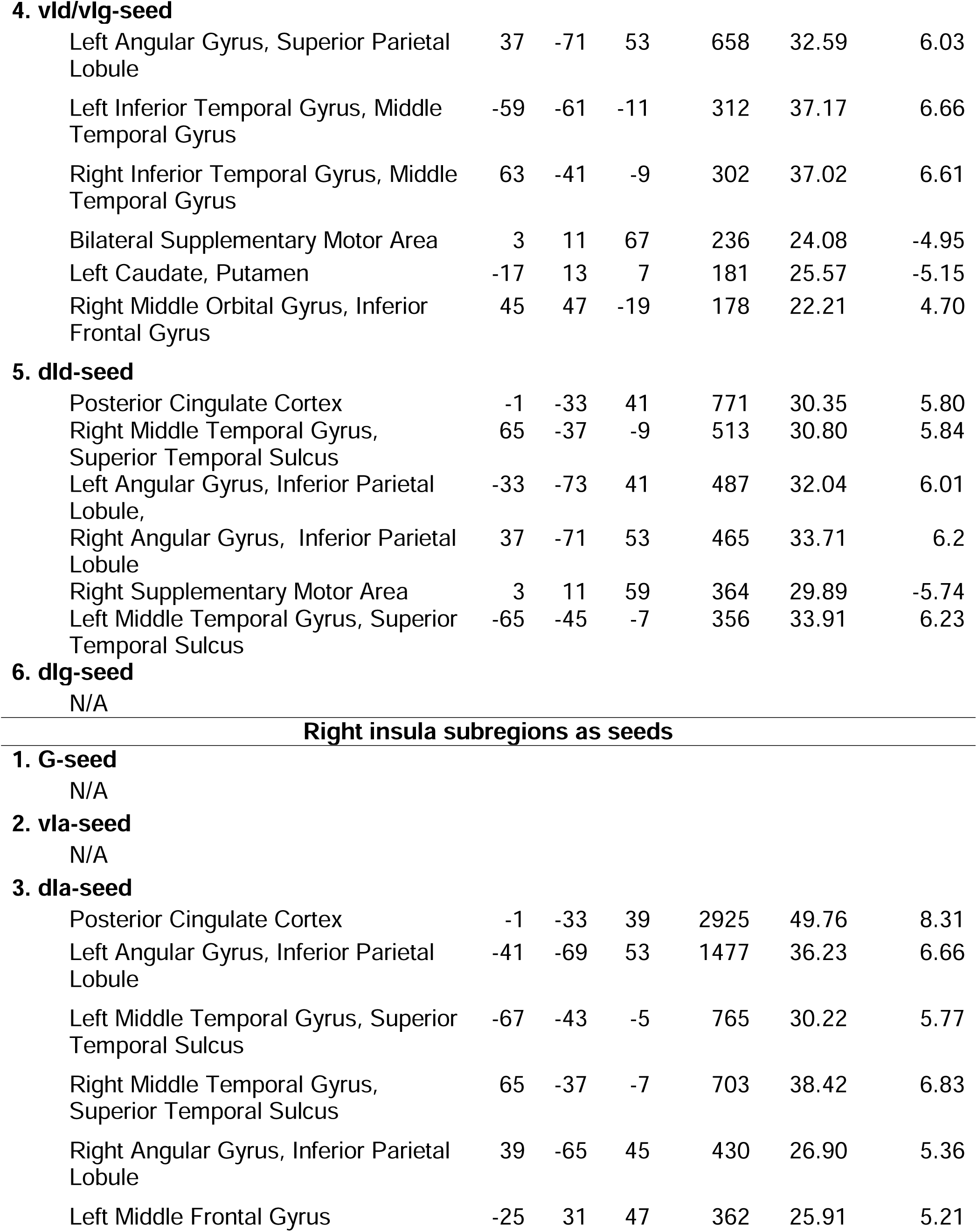
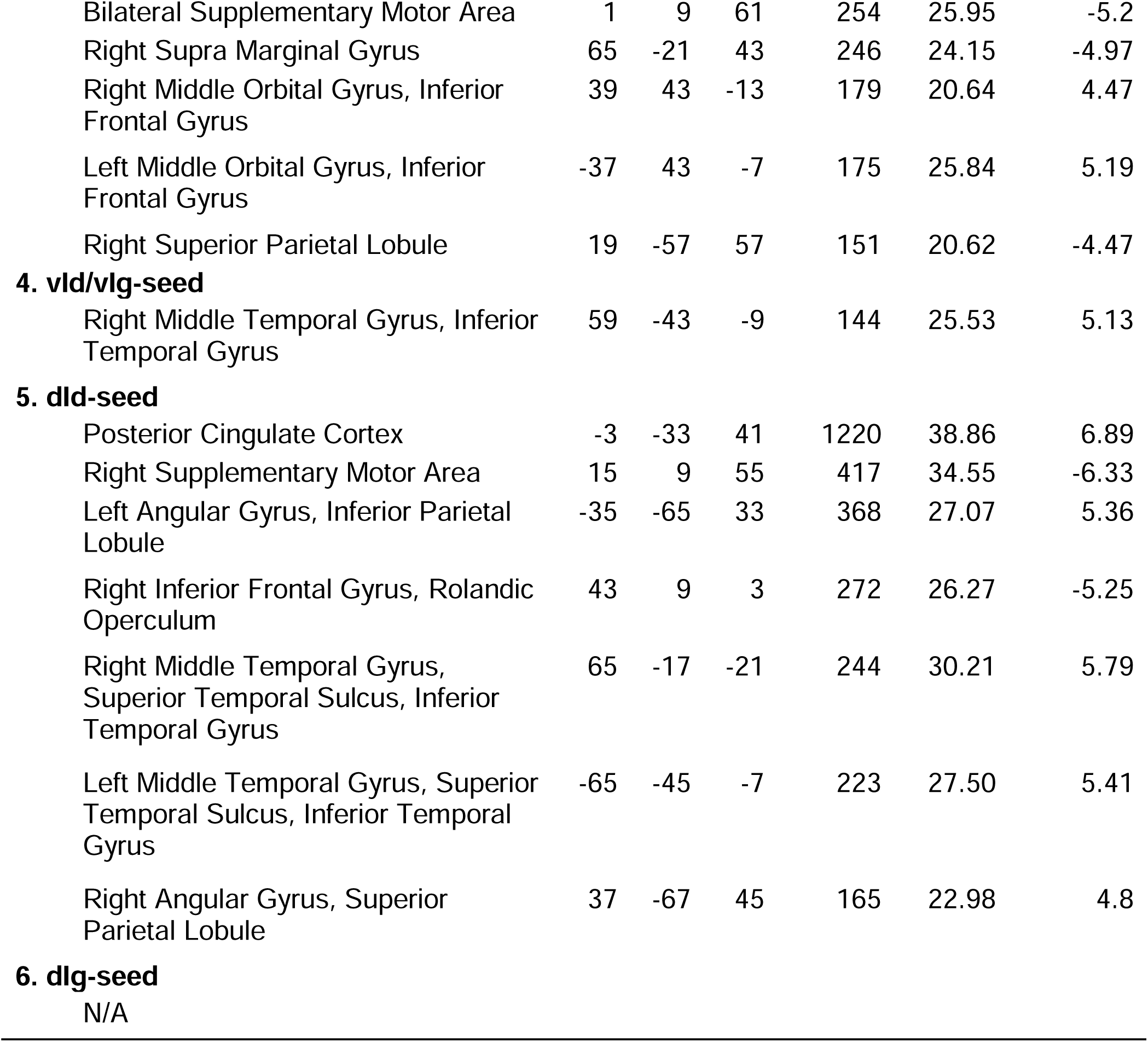
Significant regions showing main effect of run (RNT-induction and Rest) from seed-to-whole brain functional connectivity analysis.

**Figure 1.**
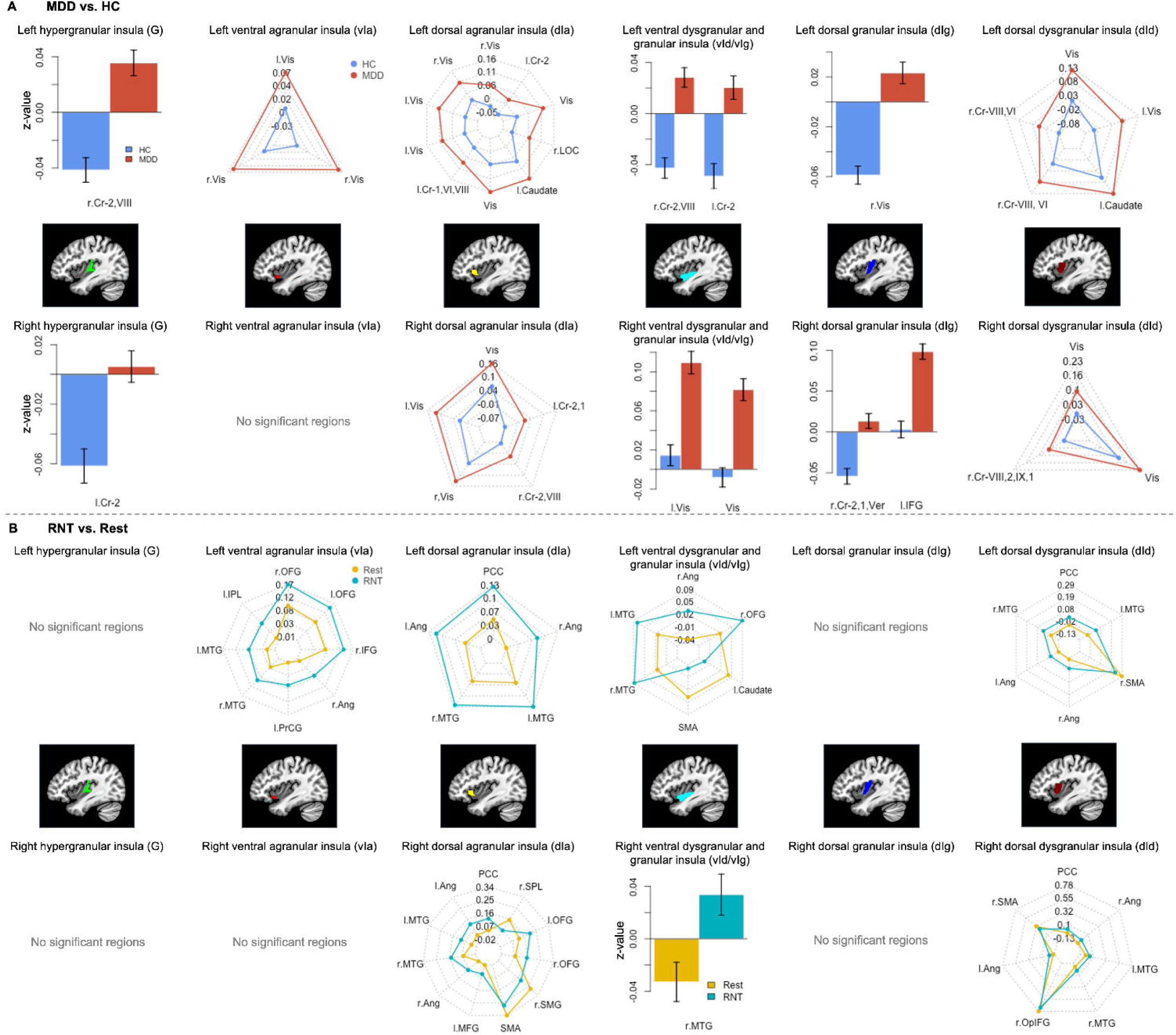
Post-hoc investigation of A) effect of diagnosis (MDD vs. HC) and B) effect of run (RNT-induction vs. Rest). Abbreviations: L - Left, r - Right, Cr - Cerebellum, Vis - Visual Area, Ver – Vermis, IFG - Inferior Frontal Gyrus, OFG - Orbital Frontal Gyrus, Ang - Angular Gyrus, PrCG – Precentral Gyrus, MTG - Middle Temporal Gyrus, IPL - Inferior Parietal Lobule, PCC - Posterior Cingulate Cortex, SMG - Supramarginal Gyrus, SPL - Superior Parietal Lobule, SMA - Supplementary Motor Area, OpIFG - Opercular part of the Inferior Frontal Gyrus.

### 3.3 Correlation between insular FC and RNT measures

Figure 2 depicts significant associations between RNT measures and FC of the insular cortex with other regions in MDD participants, as well as HC participants. Consistent with our findings in increased insular FC during RNT-induction relative to the resting-state, among individuals with MDD, higher trait-RNT was positively associated with increased FC between the right dorsal anterior and middle insula, regions in the DMN (including the posterior cingulate cortex and middle temporal gyrus), and regions in the salience network (SN) (including the orbital frontal gyrus). Moreover, greater state-RNT scores during RNT-induction, compared to resting-state, were positively correlated with increased FC in similar insular regions and the bilateral angular gyrus, as well as the right middle temporal gyrus (Figure 2). On the other hand, higher trait-RNT was negatively correlated with increased insular FC between the left anterior insula and the inferior parietal lobule in individuals with MDD, although this FC showed an increased main effect of RNT-induction (**Table 3 and Figure 1**).

**Figure 2.**
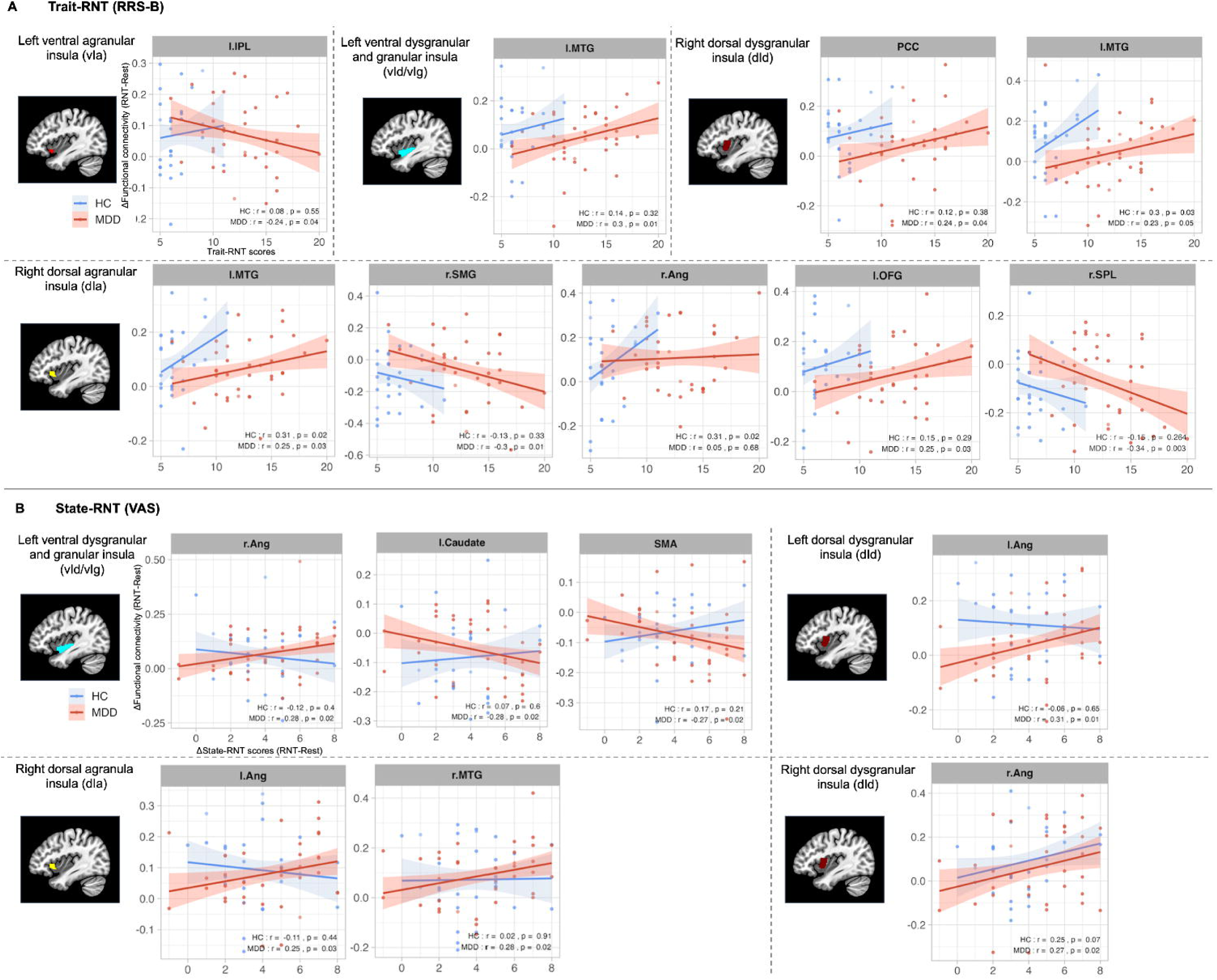
Scatter plots and correlation between insular-cortical functional connectivity (FC) and RNT measures. A) Correlation of trait-RNT as measured by the Ruminative Response Scale-Brooding subscale (RRS-B) before the scan (x-axis) with changes in FC during RNT-induction scan compared to the Rest scan (y-axis). B) Correlation of changes in state-RNT as measured by the Visual Analogue Scale (VAS) during RNT- induction scan compared to the Rest scan (x-axis) with changes in FC during RNT- induction scan compared to Rest scan (y-axis). Abbreviations: L - Left, R - Right, IPL - Inferior Parietal Lobule, MTG - Middle Temporal Gyrus, PCC - Posterior Cingulate Cortex, SMG - Supramarginal Gyrus, Ang - Angular Gyrus, OFG - Orbital Frontal Gyrus, SPL - Superior Parietal Lobule, SMA - Supplementary Motor Area.

## 4. Discussion

This study investigated the hypothesis that individuals with MDD would demonstrate a greater increase in FC between the insular cortex and other cortical (including cerebellar) regions during RNT-induction compared to resting-state. We also predicted that functional changes would be more pronounced in MDD, as compared with HC individuals. We observed three main findings during our research by which our hypothesis was partially supported. First, contrary to our hypothesis, there was no statistically significant diagnosis-by-run interaction in insular FC, indicating that changes in FC during RNT-induction are not significantly different in individuals with MDD compared to HC individuals. Second, FC between insular and cerebellar cortices was higher in individuals with MDD compared to the HC group. Third, overall, FC between insular and other cortical regions increased during RNT-induction compared to resting-state data.

Altogether, these findings support the hypothesis that the visceral control and higher-order cognitive processing changes underlie RNT intensity (Tsuchiyagaito *et al*., 2022). These findings also reflect that insular-cortical FC was stronger during RNT-induction compared to resting-state. However, our results did not demonstrate a significant difference in FC alterations during RNT-induction between the MDD and HC participants.

### 4.1 Insular Connectivity in MDD

The observed higher level in FC between the anterior, middle, and posterior insula and the cerebellum in MDD participants, as compared to healthy controls, aligns with emerging literature that emphasizes the critical role of insular alterations in emotional regulation and the pathophysiology of depression (Habas, 2021, Misaki *et al*., 2023, Pierce *et al*., 2023, Sliz and Hayley, 2012). Moreover, these increased connections between the insula and the cerebellum indicate that the cerebellum has a significant role in depression. The increased FC between the insula and the cerebellum that was observed in our research broadens our understanding of how these brain structures function independently, as well as with one another, to conceptualize and regulate emotions.

The insula, known for integrating somatosensory, affective, and cognitive information (Sliz and Hayley, 2012), may be crucial in maintaining the heightened state of the negative self-focus aspect of RNT. Prior neuroimaging research has associated emotional recalling/remembrance and other cognitively demanding, emotional tasks with increased insular activity (Phan *et al*., 2002). In this context, our observation of increased FC between the insula and other brain regions during RNT-induction tasks is not surprising. Furthermore, this suggests that the trained-regulation of insular activity, or decreasing FC between the bilateral insula and other areas such as the STS, the parietal cortices, or the posterior cingulate cortex (PCC), may reduce state-RNT symptoms. For example, the emergence of focused deep brain neuromodulation or real-time fMRI neurofeedback in FC literature prompt deeper exploration of these brain activity regulation methods as a means to improve RNT in depression.

The cerebellum’s role in cognitive and emotional processing, traditionally recognized in the last two decades (Pierce *et al*., 2023, Rudolph *et al*., 2023), appears to be particularly significant in the context of mood disorders. Previous resting-state static and dynamic FC research identifies the cerebellum as having a vital role in emotional processing and executive functioning through its connections to the executive network (EN), the DMN, the salience network (SN), the insula, and multiple brain cortical hubs associated with emotion regulation (Habas, 2021). These connections suggest that the cerebellum is involved in frequent, multimodal collaboration with other crucial brain regions and networks to undertake the multifaceted nature of human emotions. Considering the cerebellum’s association with emotion regulation and other key networks, the cerebellum may also be an ROI worth investigating in future FC studies involving clinically depressed populations.

### 4.2 Alterations During RNT Induction

The augmentation of FC during RNT-induction between insular regions and areas, such as the prefrontal and parietal cortices, posterior cingulate cortex, medial temporal gyrus, and the STS, is particularly noteworthy. These regions are implicated in a wide range of processes, from self-referential thought to emotional processing and memory retrieval. The increased connectivity that was noticed during RNT-induction in our work suggests a heightened state of neural coordination in these networks, potentially underpinning the ruminative process.

Foregoing studies have presented similar results, demonstrating significant transformations in FC that occur in state-RNT. In another mood-induction study, researchers found that increased connectivity between the DMN and the fronto-parietal network (FPN), along with decreased connectivity between the SN and the FPN, are both associated with increased RNT after experiencing sadness (Lydon-Staley *et al*., 2019). The changes in RNT-induced FC that were observed during our research, particularly with the MDD sample population, were congruent to the findings of their research. In these types of RNT-induction studies, a variety of key networks and brain regions can be observed at play in emotion regulation, many of which may serve as potential targets for interventions and future research aimed at reducing trait- and/or state- RNT symptoms.

### 4.3 Correlation between insular FC and trait- and state-RNT scores

The correlation of both trait- and state-RNT scores with increased FC in specific brain regions, particularly in MDD patients as reported herein, suggests that FC could be a potential biomarker for RNT severity in clinical settings. Specifically, trait-RNT scores were associated with the increased insular FC of several key regions in the DMN and orbitofrontal gyrus, which are implicated in self-referential and emotional processing (Northoff *et al*., 2006, REMPEL-CLOWER, 2007). This association highlights the neural correlates of a general propensity to engage in RNT, reflecting a stable, trait-like aspect of cognitive processing in individuals. In contrast, state-RNT scores were associated with FC between the insula and the angular gyrus, as well as the right medial temporal gyrus, during experimentally induced RNT. Changes in state-RNT ratings indicate how participants engaged with RNT during the experimental induction relative to the resting-state. The association with increased FC in these regions suggests that the acute induction of RNT may engage neural circuits related to memory, conceptual processing (Deen *et al*., 2015, Humphreys *et al*., 2021, Ramanan *et al*., 2018, Seghier, 2013), and the integration of emotional and sensory information (Craig, 2009). This distinction underlines the dynamic nature of RNT, where state-dependent increases in RNT were correlated with immediate neural responses, differentiating it from the more static trait-RNT. Such findings illustrate the complex neural underpinnings of RNT, supporting the idea that different facets of RNT are potentially supported by different neural networks, as reported in prior studies (Rosenbaum *et al*., 2017, Tsuchiyagaito *et al*., 2023a). However, we would caution against any definitive conclusions based on correlation analysis due to the exploratory nature of this analysis.

## 5. Limitations and Future Directions

While our findings contribute significantly to the understanding of RNT in MDD, several limitations, such as the small sample size, must be acknowledged. Longitudinal studies, or interventional studies using emerging neuromodulation methods to noninvasively modulate the large-scale circuits described herein (Philip and Arulpragasam, 2023), could help to establish a causative role of neural alterations in RNT.

Moreover, preceding research by our group and others has suggested that RNT is a transdiagnostic occurrence, as it is a usual feature in individuals with generalized anxiety disorder (GAD) and obsessive-compulsive disorder (OCD) (Wahl *et al*., 2019). Given the comorbidity of these disorders, it may be worth conducting a similar investigation that explores FC developments and trait-/state-RNT with participants from GAD and OCD populations.

## 6. Conclusion

The findings of our study underscore the importance of insular connectivity in the neural systems underlying RNT in MDD. Individuals with MDD exhibit distinct functional connectivity patterns between the insula and the cerebellum, highlighting a neural circuit that may contribute to the persistence and intensity of RNT. In addition, both MDD and healthy control participants show increased insular connectivity with key brain regions, including the bilateral prefrontal cortices, parietal lobes, posterior cingulate cortex, and medial temporal gyrus, during RNT-induction compared to resting-state. This suggests that the insula is part of a broader network that becomes more engaged during active RNT, facilitating the integration of emotional and cognitive aspects of negative self-related thoughts. Moreover, higher trait-RNT in MDD participants was associated with increased connectivity between the insula and regions within the DMN and SN, indicating that persistent negative thinking is linked to specific insular connectivity patterns involving self-referential processing and emotional salience. These differential connectivity patterns, including regions where higher trait-RNT is negatively correlated with increased insular connectivity, may serve as neural markers for the intensity of RNT.

Taken together, our findings highlight the critical role of insular connectivity and its interactions with other brain regions in the manifestation of RNT in MDD, providing a foundation for the development of targeted neuromodulatory interventions to alleviate this symptom in depression. This is in line with emerging neuromodulation techniques with anatomical specificity (Mehić *et al*., 2014, Siddiqi *et al*., 2020) that can be used to modulate this circuitry.

## Data Availability

External researchers can make written requests for data sharing. Requests will be assessed on a case-by-case basis in consultation with the lead and coinvestigators.

## Author Contributions

Conceptualization: Landon S Edwards and Aki Tsuchiyagaito; methodology and formal analysis: Landon S Edwards, Masaya Misaki, Aki Tsuchiyagaito; writing – original draft: : Landon S Edwards, Salvador M Guinjoan, and Aki Tsuchiyagaito; writing – review and editing: Saampras Ganesan, Jolene Tay, Eli S Elliott, Masaya Misaki, Martin P Paulus, Salvador M Guinjoan, Evan J White; resources: Masaya Misaki and Martin P. Paulus; supervision: Martin P. Paulus, and Salvador M. Guinjoan; funding acquisition: Martin P. Paulus.

## Role of the Funding Sources

This work has been supported in part by the National Institute of General Medical Sciences Center Grant Award Number, P20GM121312 and the Laureate Institute for Brain Research. The content is solely the responsibility of the authors and does not necessarily represent the official views of the National Institutes of Health.

## Acknowledgments

We would like to express our appreciation to CoBRE NeuroMap Investigators at LIBR and all the research participants. We acknowledge the contributions of Sahib S. Khalsa, M.D., Ph.D., Tim Collins, Dara Crittenden, Amy Peterson, Megan Cole, Lisa Kinyon, Lindsey Bailey, Courtney Boone, Natosha Markham, Lisa Rillo, Angela Yakshin, and the LIBR Assessment Team for diagnostic assessments and data collection, and Julie Arterbury, Leslie Walker, Amy Ginn, Bill Alden, Julie DiCarlo, and Greg Hammond for helping with MRI scanning. The authors acknowledge Jerzy Bodurka, Ph.D. (1964– 2021) for his intellectual and scientific contributions to the establishment of the EEG, structural and functional MRI, and neurofeedback processes that provided the foundation for the data collection, analysis, and interpretation of findings for the present work.

## Conflict of Interest Disclosure

Dr. Martin P. Paulus is an advisor to Spring Care, Inc., a behavioral health startup, and he has received royalties for an article about methamphetamine in UpToDate. Dr. Martin P. Paulus has a consulting agreement with and receives compensation from F. Hoffmann-La Roche Ltd. The other authors report no financial relationships with commercial interests related to the present study.

## Notes

### Competing Interest Statement

Dr. Martin P. Paulus is an advisor to Spring Care, Inc., a behavioral health startup, he has received royalties for an article about methamphetamine in UpToDate. Dr. . Martin P. Paulus has a consulting agreement with and receives compensation from F. Hoffmann-La Roche Ltd. The other authors report no financial relationships with commercial interests related to the present study.

### Clinical Trial

NCT04941066

### Author Declarations

The study protocol was reviewed and approved by the WCG IRB (https://www.wcgirb.com) (IRB Tracking Number 20210286).

