## Supplemental materials for "Increased Insular Functional Connectivity During Repetitive Negative Thinking in Major Depression and Healthy Volunteers"

### 1. Supplementary figures

**Figure S1**

**Insula subregions defined based on the Brainnetome atlas.**

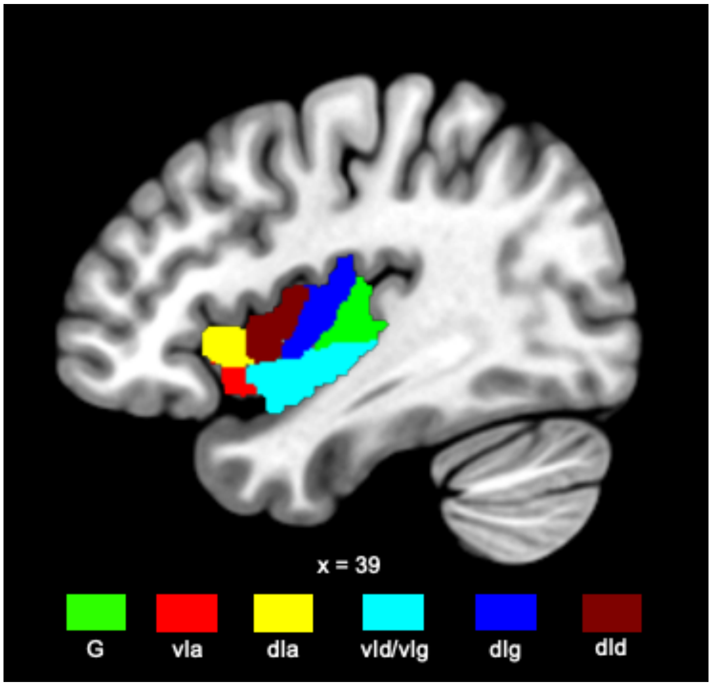

Note. G, hypergranular insula; vIa, ventral agranular insula; dIa, dorsal agranular insula; vId/vIg, ventral dysgranular and granular insula; dIg, dorsal granular insula; dId, dorsal dysgranular insula

**Figure S2**

Functional connectivity during RNT compared to Rest in individuals with MDD, relative to HC (left insula subregions).

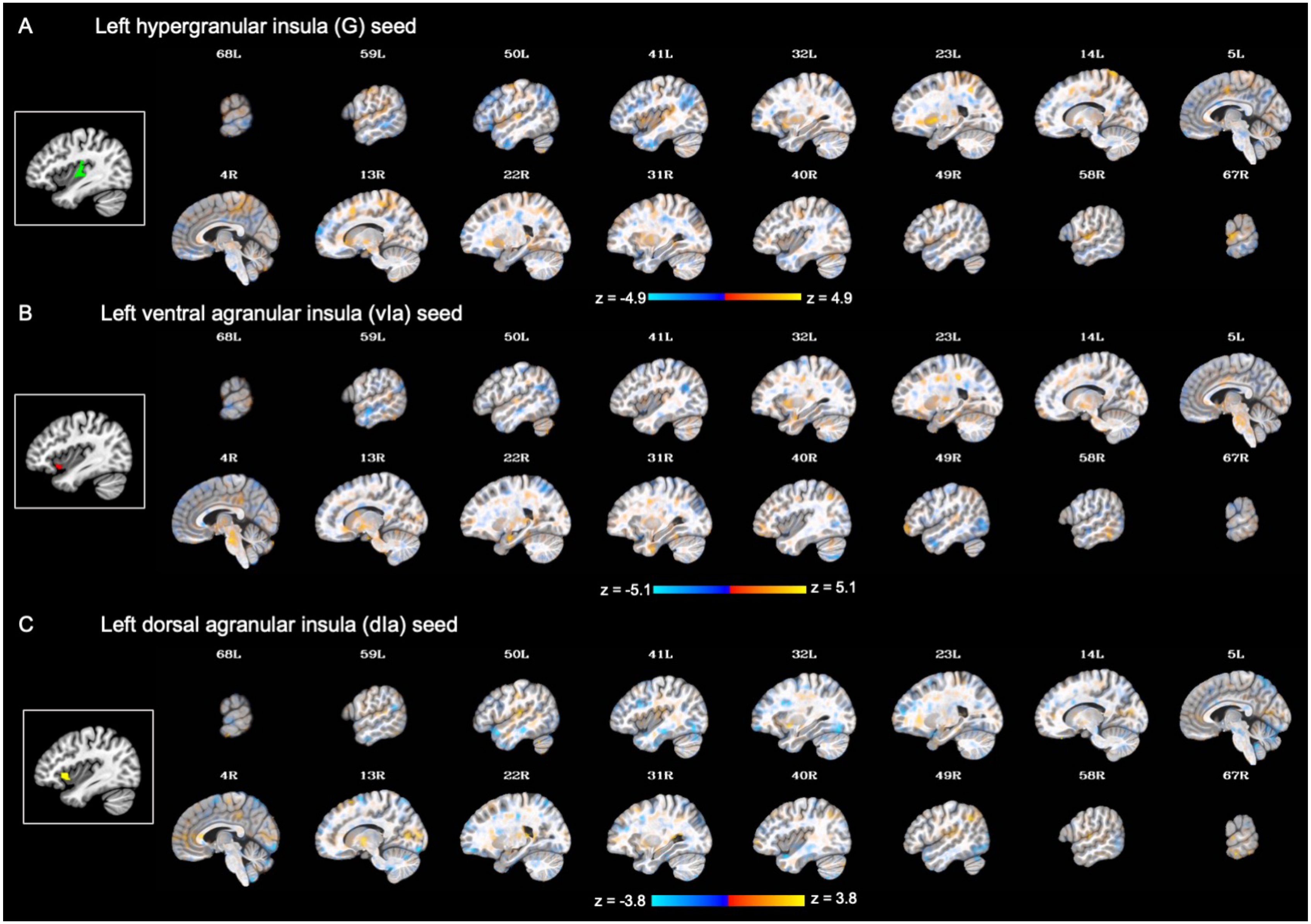

**Figure S2 (Continued)**

Functional connectivity during RNT compared to Rest in individuals with MDD, relative to HC (left insula subregions).

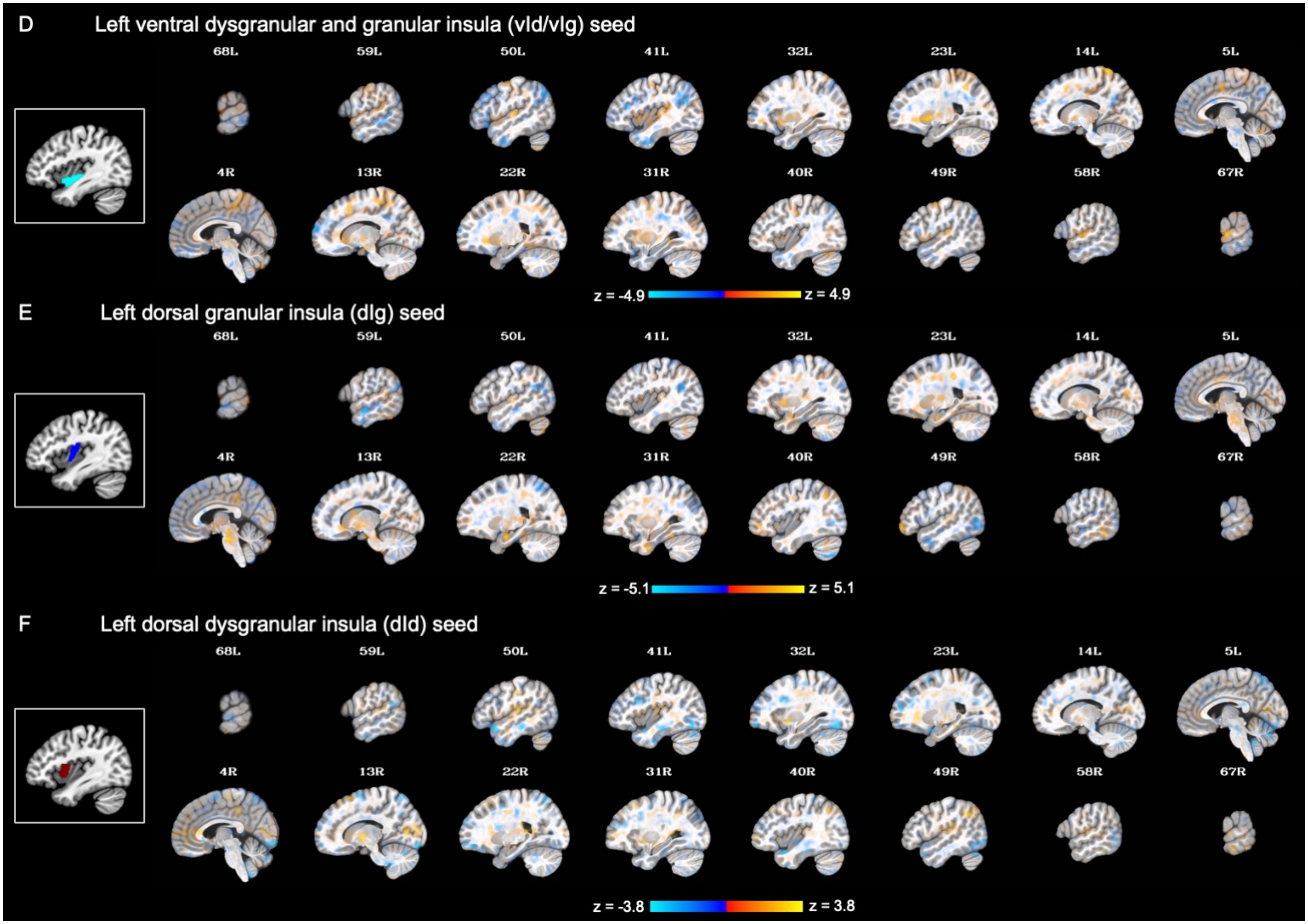

Note. A translucent statistical threshold set at $p < 0.001$ is applied, without cluster-extent correction. Areas exhibit increasing transparency corresponding to decreasing statistical significance (Taylor et al., 2023). Each seed region is displayed on the left side of its corresponding statistical map, framed with a white square outline.

**Figure S3**

Functional connectivity during RNT compared to Rest in individuals with MDD, relative to HC (right insula subregions).

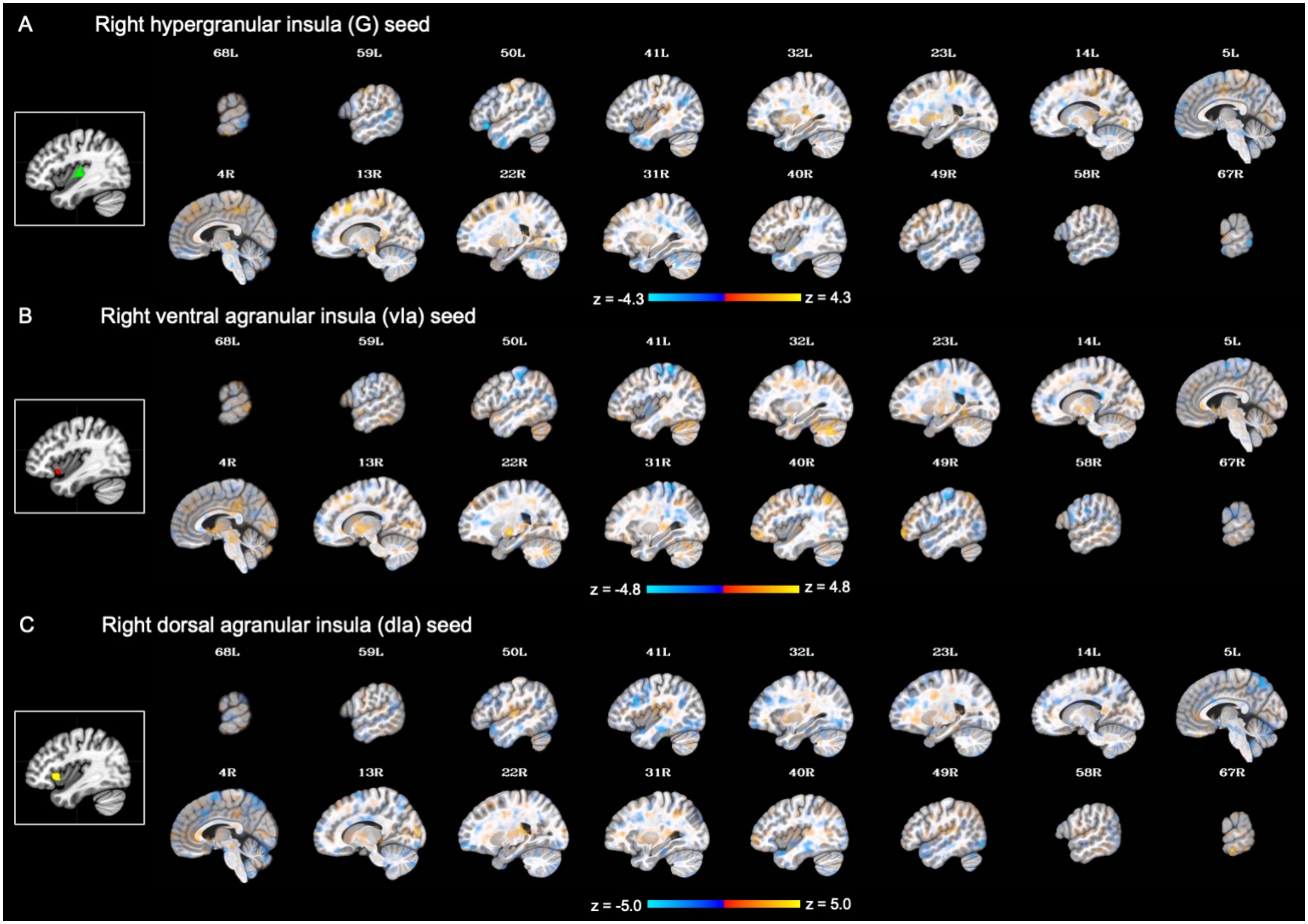

**Figure S3 (Continued)**

Functional connectivity during RNT compared to Rest in individuals with MDD, relative to HC (right insula subregions).

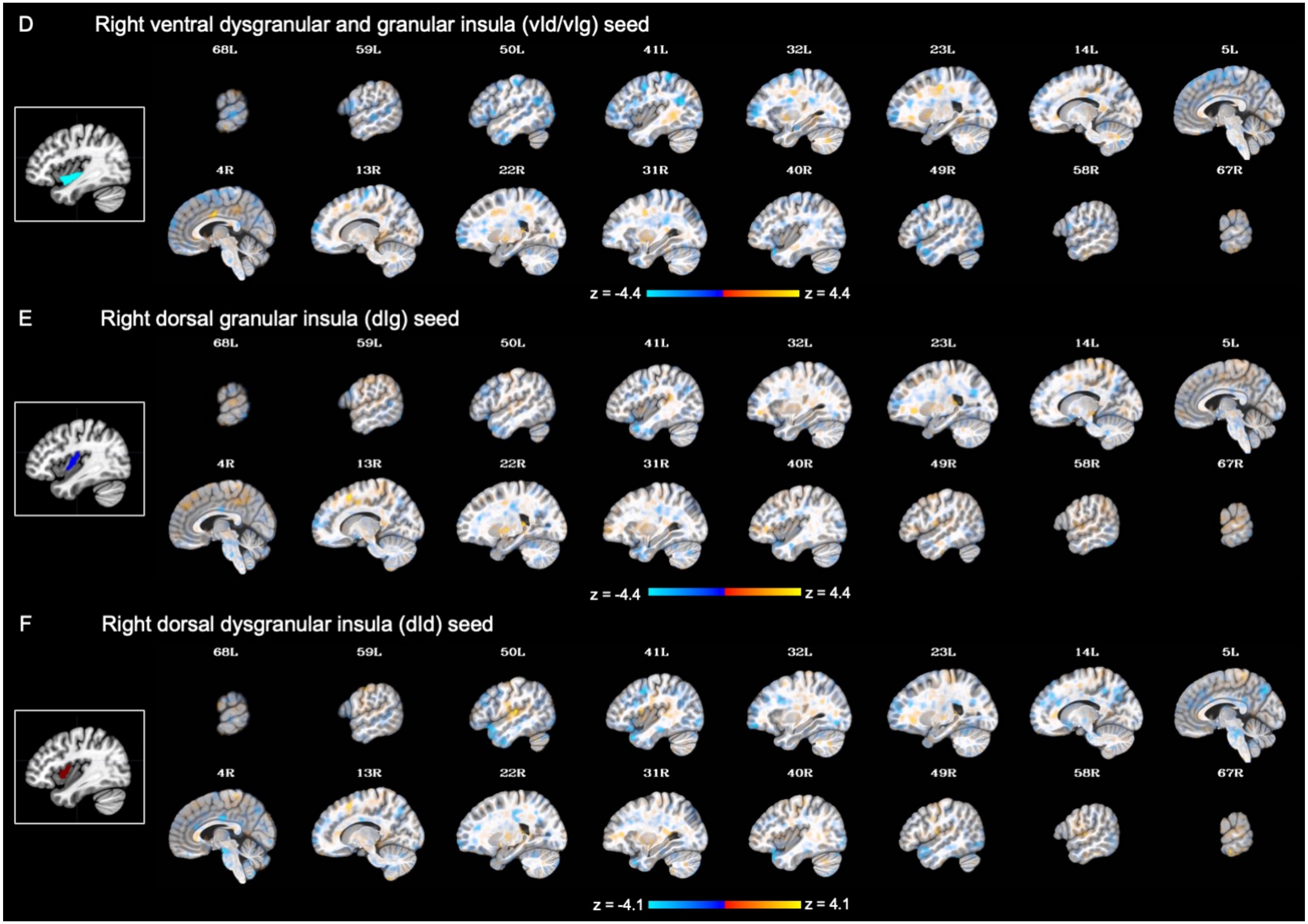

Note. A translucent statistical threshold set at $p < 0.001$ is applied, without cluster-extent correction. Areas exhibit increasing transparency corresponding to decreasing statistical significance (Taylor et al., 2023). Each seed region is displayed on the left side of its corresponding statistical map, framed with a white square outline.

**Figure S4**

Effect of diagnosis (MDD vs. HC) on seed-to-whole brain functional connectivity (left insula subregions as seeds).

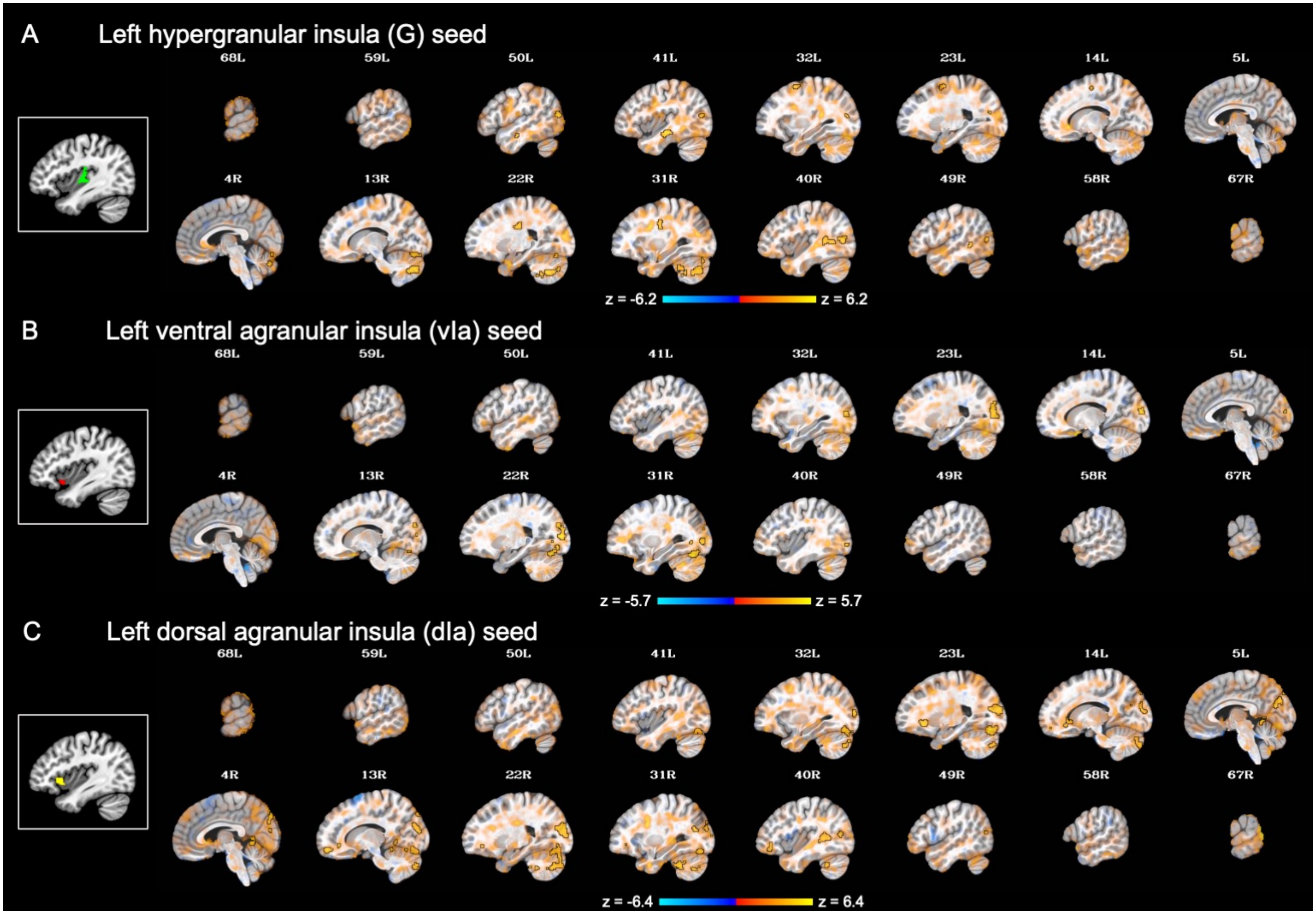

**Figure S4 (Continued)**

Effect of diagnosis (MDD vs. HC) on seed-to-whole brain functional connectivity (left insula subregions as seeds).

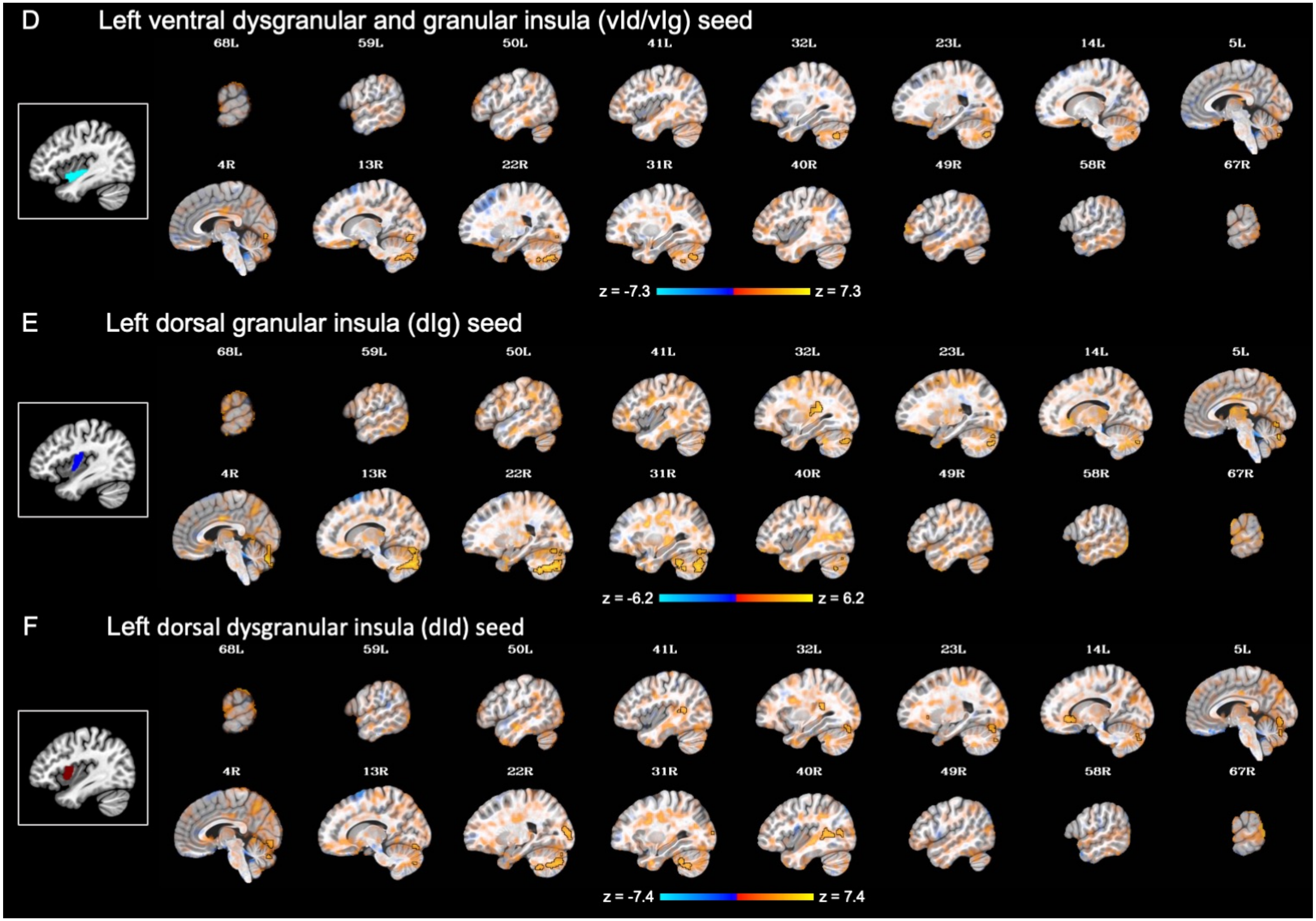

Note. A translucent statistical threshold is applied at $p < 0.001$, with cluster-extent correction at $k > 143$. Suprathreshold areas are emphasized with opacity and black outlines, while sub-threshold areas gradually become more transparent as their statistical significance decreases (Taylor et al., 2023).

**Figure S5**

Effect of diagnosis (MDD vs. HC) on seed-to-whole brain functional connectivity (right insula subregions as seeds).

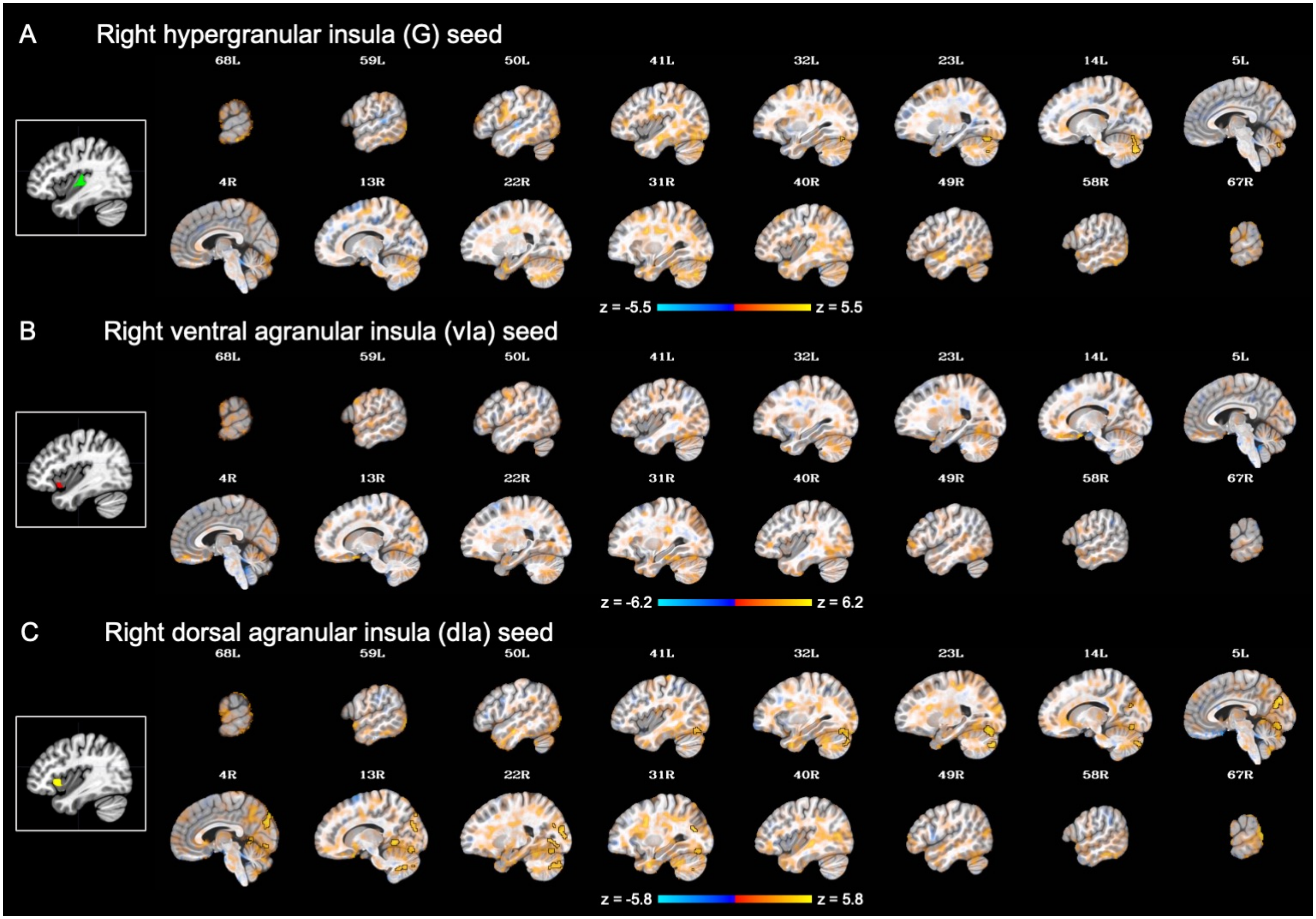

**Figure S5 (Continued)**

Effect of diagnosis (MDD vs. HC) on seed-to-whole brain functional connectivity (right insula subregions as seeds).

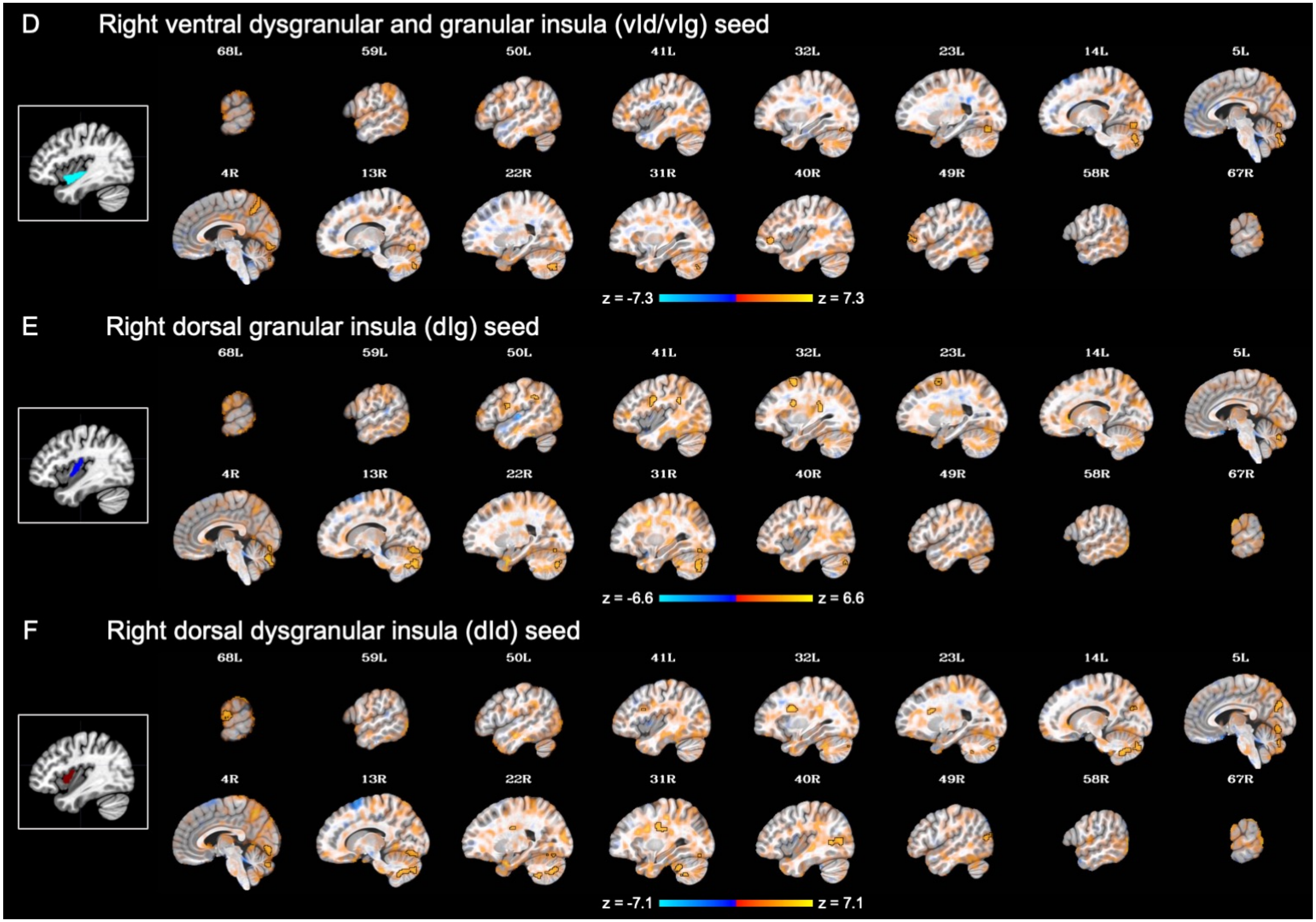

Note. A translucent statistical threshold is applied at $p < 0.001$, with cluster-extent correction at $k > 143$. Suprathreshold areas are emphasized with opacity and black outlines, while sub-threshold areas gradually become more transparent as their statistical significance decreases (Taylor et al., 2023).

**Figure S6**

Effect of run (RNT vs. Rest) on seed-to-whole brain functional connectivity (left insula subregions as seeds).

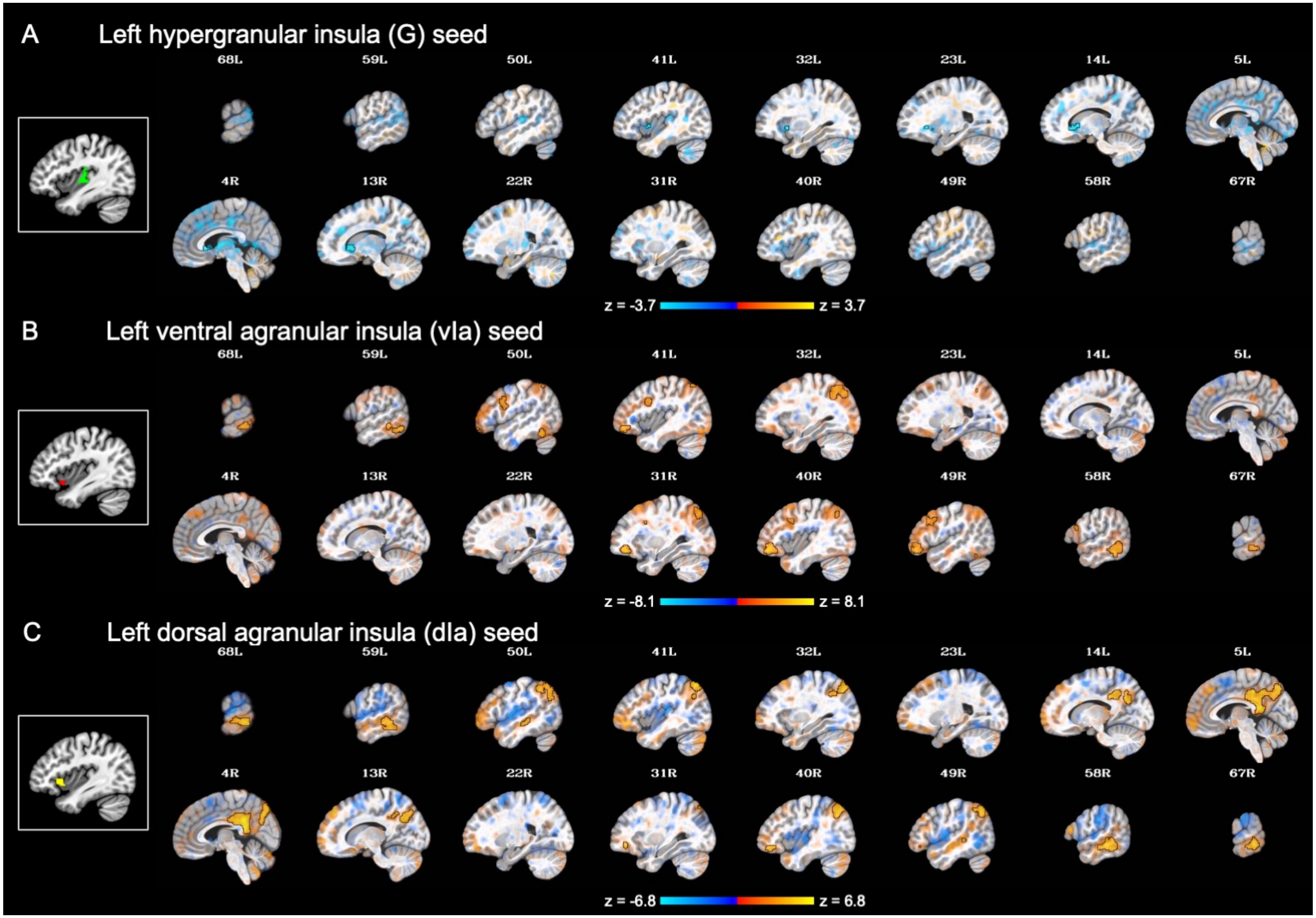

**Figure S6 (Continued)**

Effect of run (RNT vs. Rest) on seed-to-whole brain functional connectivity (left insula subregions as seeds).

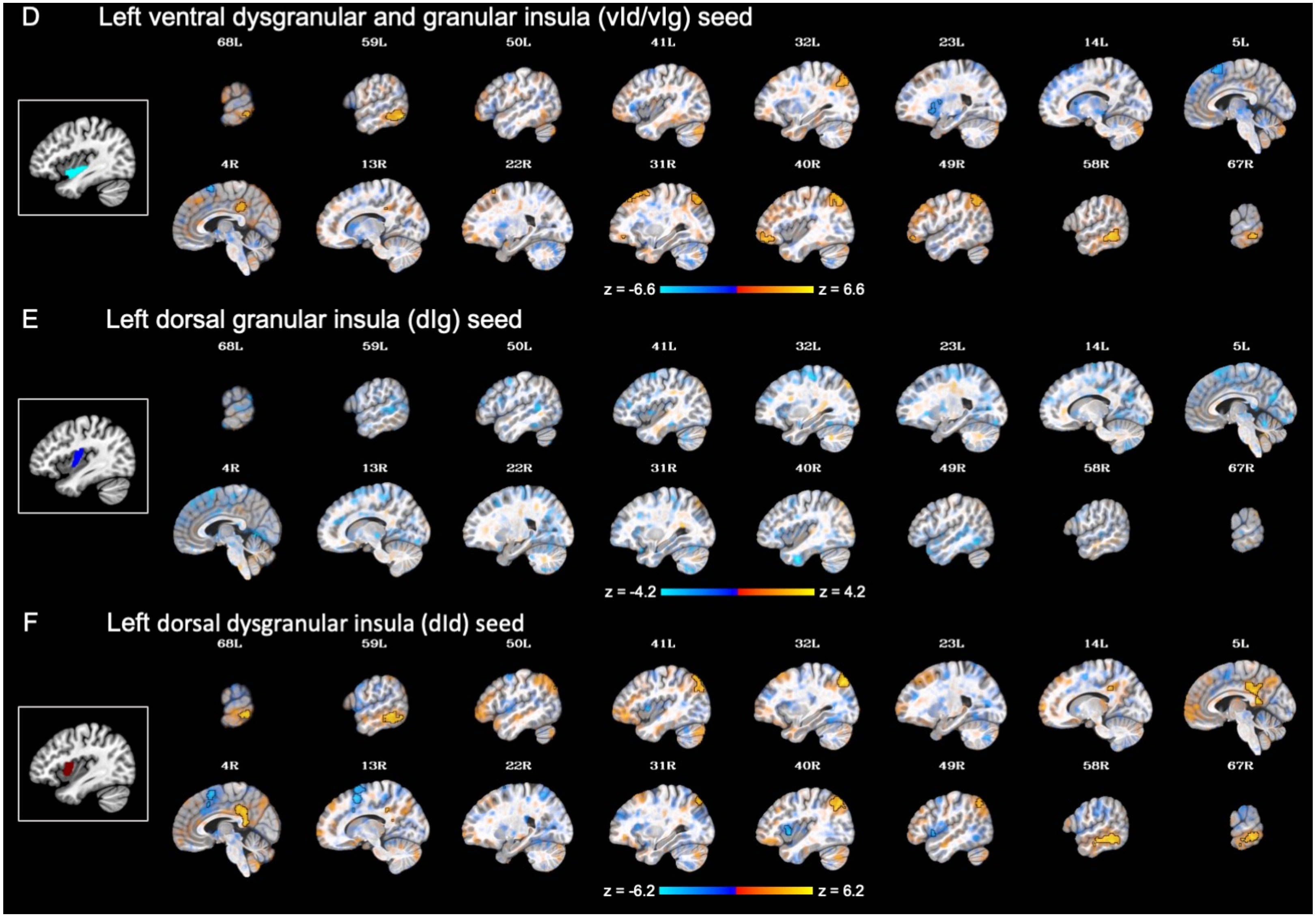

Note. A translucent statistical threshold is applied at $p < 0.001$, with cluster-extent correction at $k > 143$. Suprathreshold areas are emphasized with opacity and black outlines, while sub-threshold areas gradually become more transparent as their statistical significance decreases (Taylor et al., 2023).

**Figure S7**

Effect of run (RNT vs. Rest) on seed-to-whole brain functional connectivity (right insula subregions as seeds).

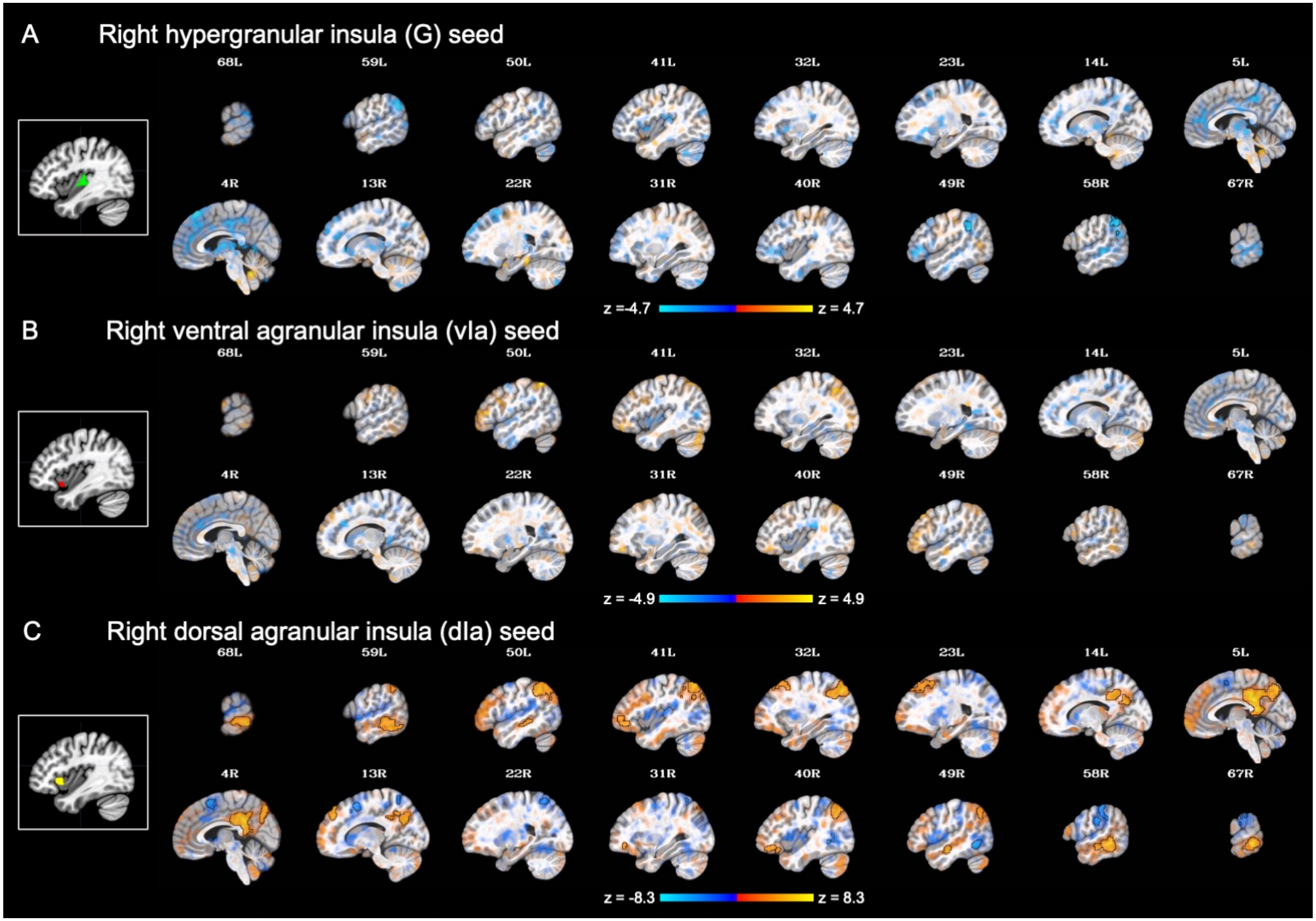

**Figure S7 (Continued)**

Effect of run (RNT vs. Rest) on seed-to-whole brain functional connectivity (right insula subregions as seeds).

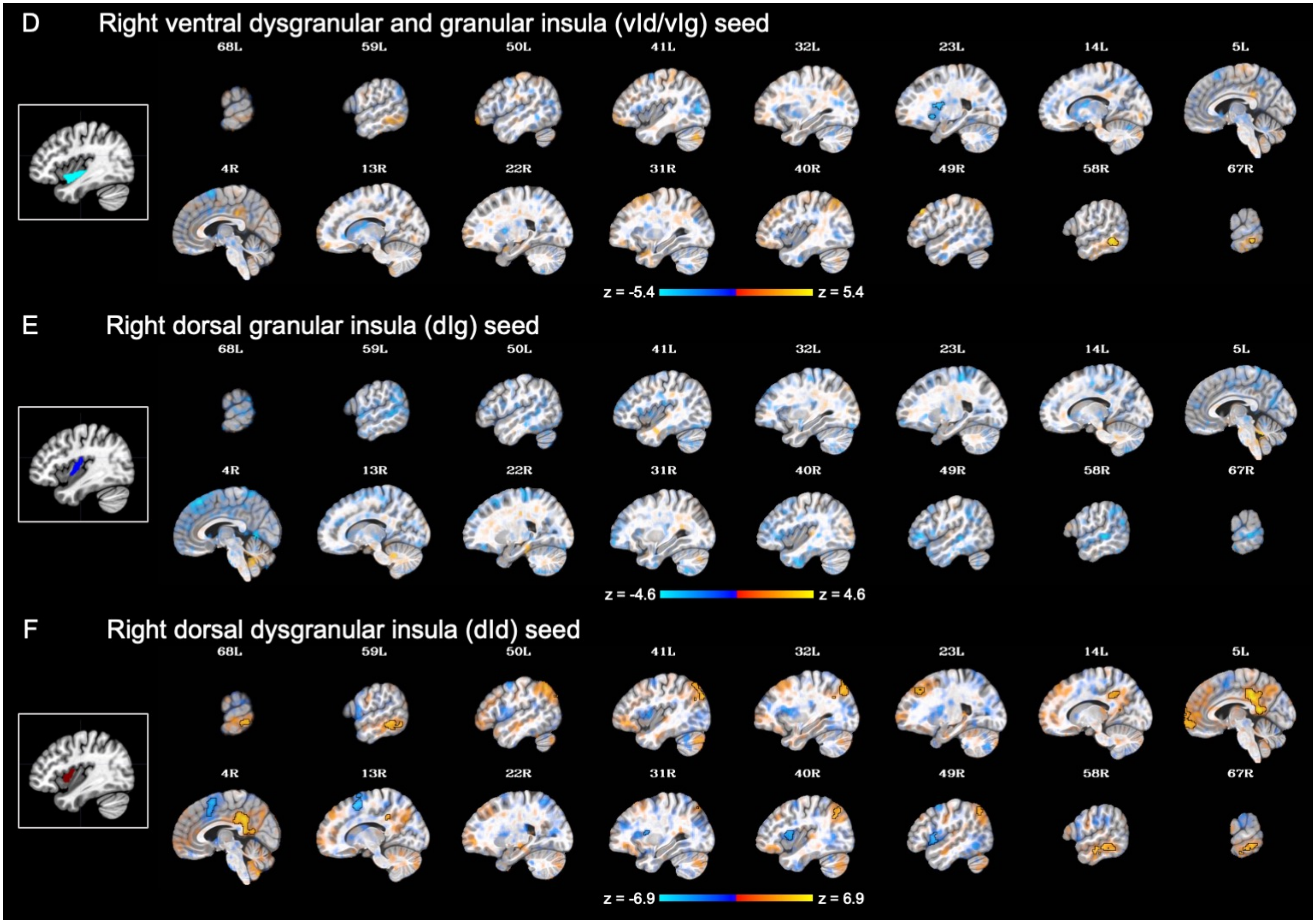

Note. A translucent statistical threshold is applied at $p < 0.001$, with cluster-extent correction at $k > 143$. Suprathreshold areas are emphasized with opacity and black outlines, while sub-threshold areas gradually become more transparent as their statistical significance decreases (Taylor et al., 2023).
